# Diabetes, glycaemic profile and risk of vitiligo: a Mendelian randomisation study

**DOI:** 10.1101/2023.10.30.23297752

**Authors:** Shucheng Hu, Yuhui Che, Jiaying Cai, Jing Guo, Jinhao Zeng

**Affiliations:** Chengdu University of Traditional Chinese Medicine, Chengdu, China; Hospital of Chengdu University of Traditional Chinese Medicine, Chengdu, China

**Keywords:** Vitiligo, Abnormal blood glucose traits, Type 1 diabetes mellitus, Type 2 diabetes mellitus, Mendelian randomization

## Abstract

**Objectives:** Previous observational studies have shown that vitiligo usually co-manifests with a variety of dysglycemic diseases, such as Type 1 diabetes mellitus(T1DM) and Type 2 diabetes mellitus(T2DM). Mendelian randomization analysis was performed to further evaluate the causal association between fasting plasma glucose, glycosylated hemoglobin(HbA1c),T1DM,T2DM and vitiligo.

**Methods:** We used aggregated genome-wide association data from the (Integrative Epidemiology Unit) IEU online database of European adults vitiligo; Glycated hemoglobin (HbA1c) data were from (IEU). Fasting blood glucose data were obtained from the European Bioinformatics Institute(EBI). T1DM and T2DM data were from FinnGen(FINN).

We used bidirectional two-sample and multivariate Mendelian randomization analyses to test whether dysglycemic measures (fasting blood glucose, HbA1c), diabetes-related measures (T1DM, T2DM) are causatively associated with vitiligo. IVW method was used as the main test method, MR-Egger, Weighted mode and Weighted median were used as supplementary methods.

**Results:** We found no statistically significant evidence to support a causal association between dysglycemic traits and vitiligo, but in the correlation analysis of diabetic traits, our data supported a positive causal association between T1DM and vitiligo (p=0.018; 95%OR:1.000(1.000-1.000)); In the follow-up multivariate MR Analysis, our results still supported this conclusion (p=0.016, 95% OR= 1.000(1.000-1.000)), and suggested that Hba1c was not a mediator of T1DM affecting the pathogenesis of vitiligo. No reverse causality was found in any of the reverse MR Analyses of dysglycemic traits and diabetic traits.

**Conclusions:** Our findings support that T1DM is a risk factor for the development of vitiligo, and this conclusion may explain why the co-presentation of T1DM and vitiligo is often seen in observational studies. Clinical use of measures related to T1DM may be a new idea for the prevention or treatment of vitiligo.

## Introdution

Vitiligo is a pigment-loss disorder with a global incidence of about 1%(1). In addition to affecting the appearance, vitiligo can also lead to the disorder of the patient’s autoimmune system. The risk of autoimmune diseases in vitiligo patients is significantly higher than that in normal people. For example, several controlled trials(2) have been conducted in East Asian and African populations, Chang etal.(3) found that the incidence of diabetes in patients with vitiligo was significantly higher than that in normal people, and believed that there was a correlation effect between vitiligo and diabetes, but the causal relationship was not clear. Such a conclusion is not confirmed only at the individual level. Existing studies (4)have shown that in the peripheral blood and skin tissue fluid of patients with vitiligo, the levels of pro-inflammatory cytokines IL-6, IL-β, TNF-α and CD8 cells are increased, while the related indicators in patients with diabetes also have an increasing trend. It is not a coincidence that the levels of inflammatory cytokines such as IL-6 and CD8 cells are often increased in the peripheral blood of patients with diabetes. At the big data analysis level, a meta-analysis(3) showed that vitiligo was significantly associated with diabetes (OR 2.515, 95% CI 1.972- 3.208; p: 0.001). However, it is not clear whether there is a causal relationship between vitiligo and diabetes.

Although a number of studies have shown that factors such as blood glucose characteristics and diabetes mellitus are associated with the occurrence of vitiligo, the current studies are mainly observational studies, and the results are susceptible to confounding factors and reverse causality. For diabetic features, the causal inference between blood glucose features and vitiligo is less convincing. The MR Method can effectively avoid the interference of confounding factors and reverse causality.

Understanding the causal relationship between vitiligo and diabetes mellitus can help us prevent or treat the occurrence of diabetes in vitiligo patients, so it is of practical significance to explore the causal relationship between vitiligo and diabetes.

Mendelian randomization (5)is a data-analysis method that uses genetic variables as instrumental variables for exposure in order to study causal associations between exposure and outcomes. It was first proposed by Katan(6) in 1986. Because the genetic alleles associated with exposure follow the Mendelian inheritance law and are randomly combined at the time of conception, MR Analysis can greatly avoid the interference of environmental factors and other diseases, thereby enhancing the inference of causal association of exposure outcomes(7). Therefore, we conducted a two-sample Mendelian randomization (TSMR) study to investigate the causal relationship between levels of fasting plasma glucose, glycosylated hemoglobin, type 1 diabetes mellitus, type 2 diabetes mellitus and vitiligo.

## Materials and Methods

### Study Design

The study adhered to Enhanced Reporting of Observational studies in epidemiology (8)- Guidelines for reporting Mendelian randomization. The study design is shown in S 1 Fig. Study design and workflow. In order to explore the causal relationship between blood glucose characteristics (including fasting blood glucose and glycosylated hemoglobin), diabetes characteristics (including type 1 diabetes, type 2 diabetes) and vitiligo, we performed a total of three MR Analyses. First, we used univariate Mendelian randomization to investigate the overall effect of blood glucose and diabetes characteristics on vitiligo outcome and performed an inverse MR Analysis to investigate the effect of vitiligo on dysglycemia and the onset of type T1 and T2DM. On this basis, multivariable MR Models were constructed to analyze the direct or independent effects of Hba1c, T1DM, and other characteristics on outcomes. The model was developed to determine the major factors affecting the prognosis of vitiligo, At present, some studies(9–11) have suggested that glycosylated hemoglobin has statistical significance in peripheral blood of vitiligo patients, and glycosylated hemoglobin is one of the indicators of type 1 diabetes.

### Data sources

Our study used publicly available GWAS data for which informed consent and ethical approval had previously been obtained. We used aggregated genome-wide association data from the (Integrative Epidemiology Unit) IEU online database of European adults vitiligo; Glycated hemoglobin (HbA1c) data were from (IEU). Fasting blood glucose data were obtained from the European Bioinformatics Institute(EBI). T1DM and T2DM data were from FinnGen(FINN).Detailed GWAS data are shown in the table below (S1 Table1).

**Supplementary Table 1.**
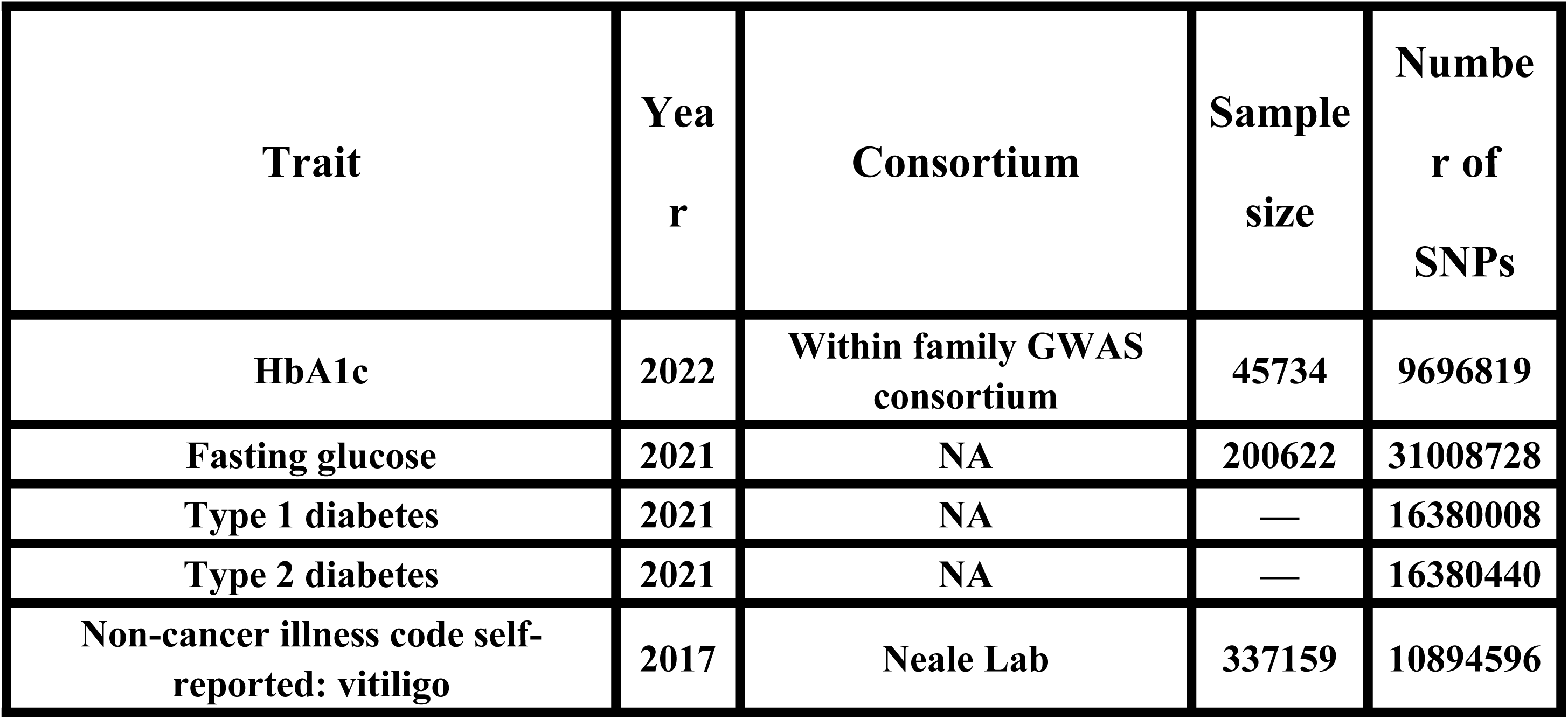
Data Source.

### Instrumental variable selection

The selection of instrumental variables generally needs to meet the following three assumptions(12):

1. Correlation hypothesis; The instrumental variable must have a strong association with the exposure (p value < 0.05).
2. Assumption of independence; Instrumental variables were not associated with confounders.
3. Assumption of exclusivity; Instrumental variables can only affect outcomes through exposure and cannot be directly correlated with outcomes.

In the univariate Mendelian randomization phase, we first tested for horizontal pleiotropy of exposures and outcomes and ensured that assumptions of independence were valid (linkage disequilibrium,LD cluster r^2^ threshold of 0.001 and window size of 10 Mb). If the IVW test results were relevant (p value < 0.05), we performed sensitivity analyses for each trait.

For multivariable Mendelian randomization, we constructed a multivariable Mendelian randomization model using type 1 diabetes mellitus, glycated hemoglobin, and vitiligo. SNPs for these traits were combined to remove confounding factors. To avoid bias caused by potentially weak instrumental variables, we used the F-statistic (calculated by the formula *F* = *beta*2/*se*2) to evaluate the strength of IV. If the F-statistic > 10, then the correlation between IV and exposure is considered strong enough to guarantee that the results of the MR Analysis are protected from weak instrumental variable bias(13).

### MR Analysis

In univariate MR Analysis, we used the IVW method as our primary test and performed sensitivity tests, such as MR-Egger(14); Weighted Median(15); Weighted Mode(16); MR-MR-PRESSO(17) was used to check for heterogeneity and horizontal pleiotropy to help inform confidence in the results. On this basis, we performed reverse MR Analysis. Specific methodological procedures were the same as for univariate MR Analysis. On the basis of univariate MR Analysis, multivariate MR Models were constructed for abnormal blood glucose, type 1 diabetes mellitus, type 2 diabetes mellitus and vitiligo. A total of 42 nSNPs were extracted, and after harmonise was performed to remove unmatchable SNPs, 32 SNPs were retained for multivariate MR Analysis. At equilibrium, the Inverse-Variance Weighted(IVW) method was still used as the main method to test the results, and in the case of positive results, we used multivariate MR-Egger (intercept), MR-PRESSO, and Weighted Mode to test for heterogeneity and horizontal pleiotropy of multivariate results, and heterogeneity, defined as variation in causal estimates of different SNPs, was used to assess heterogeneity of different SNPs in multivariate MR.

## Results

The data selected for this study were all from the European population, and the number of instrumental variables and their correlations are shown in S2-S3 Tables. In univariate MR Analysis, F statistics for all SNPs were > 10, a result that suggests no potential causal bias in the data.

**Supplementary Table 2.**
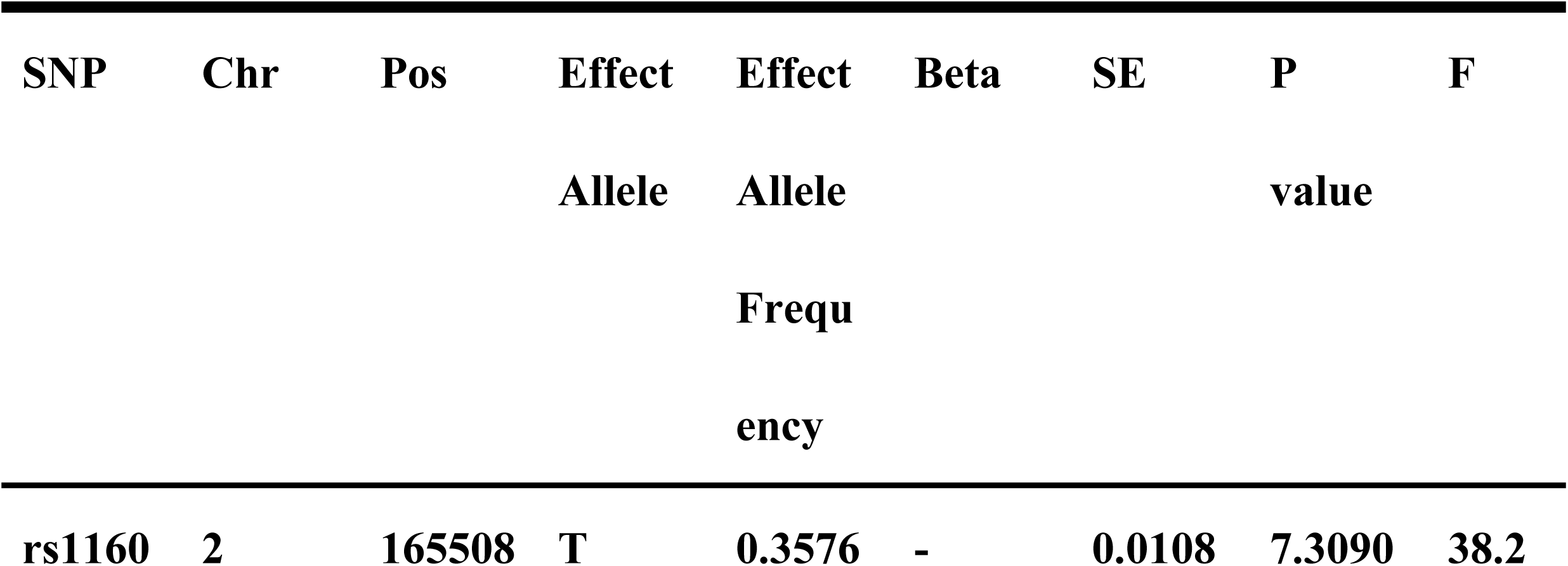
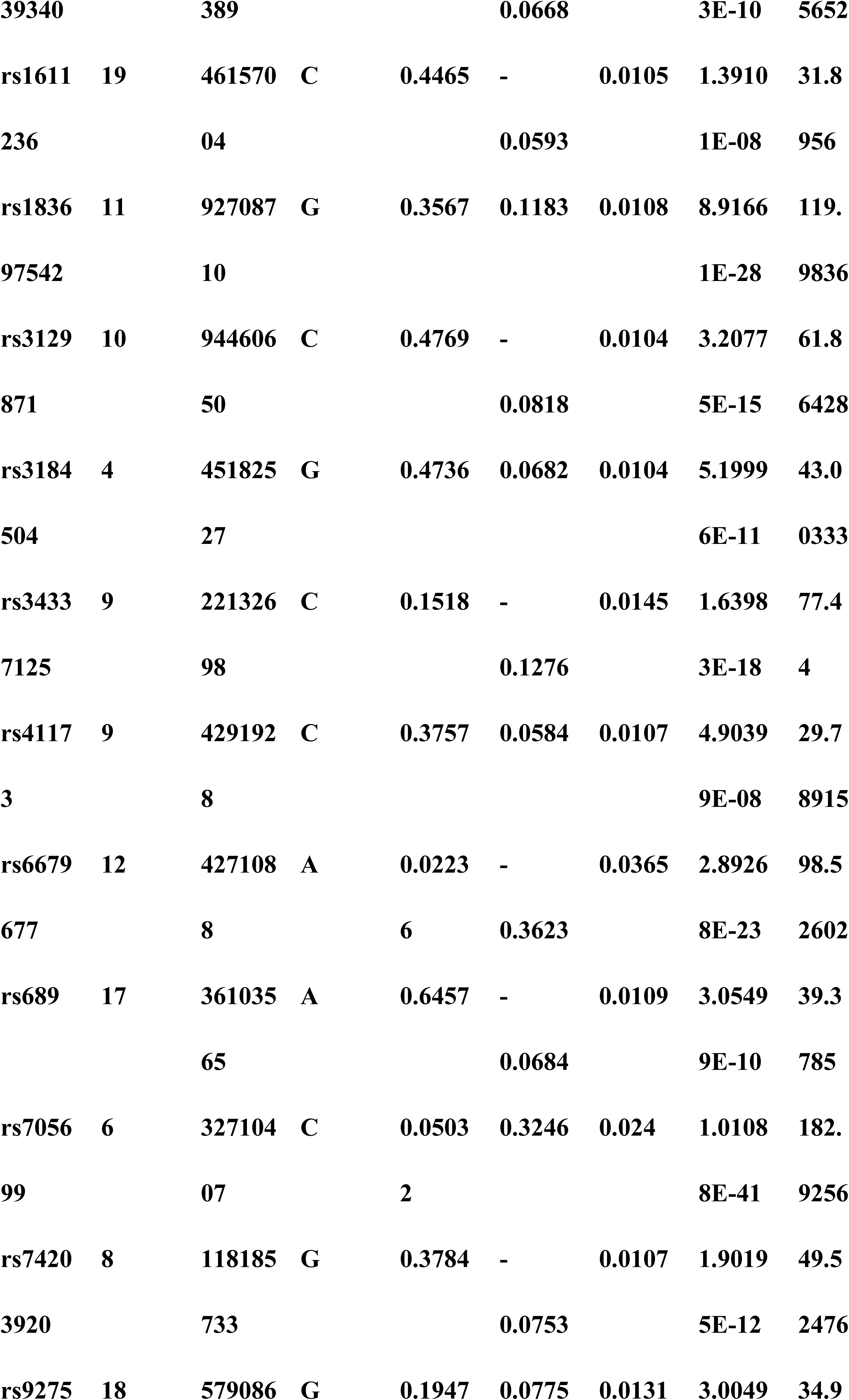
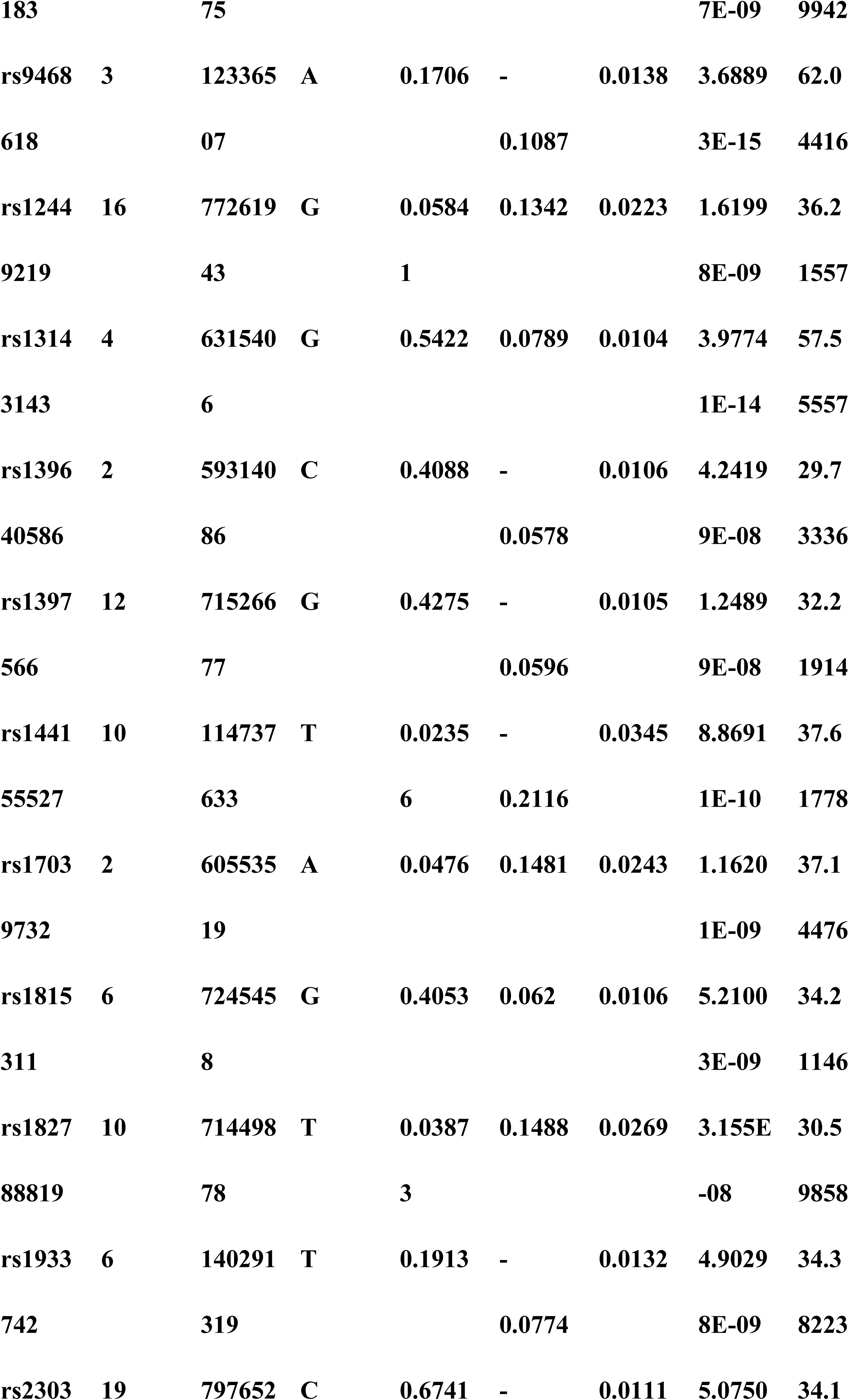
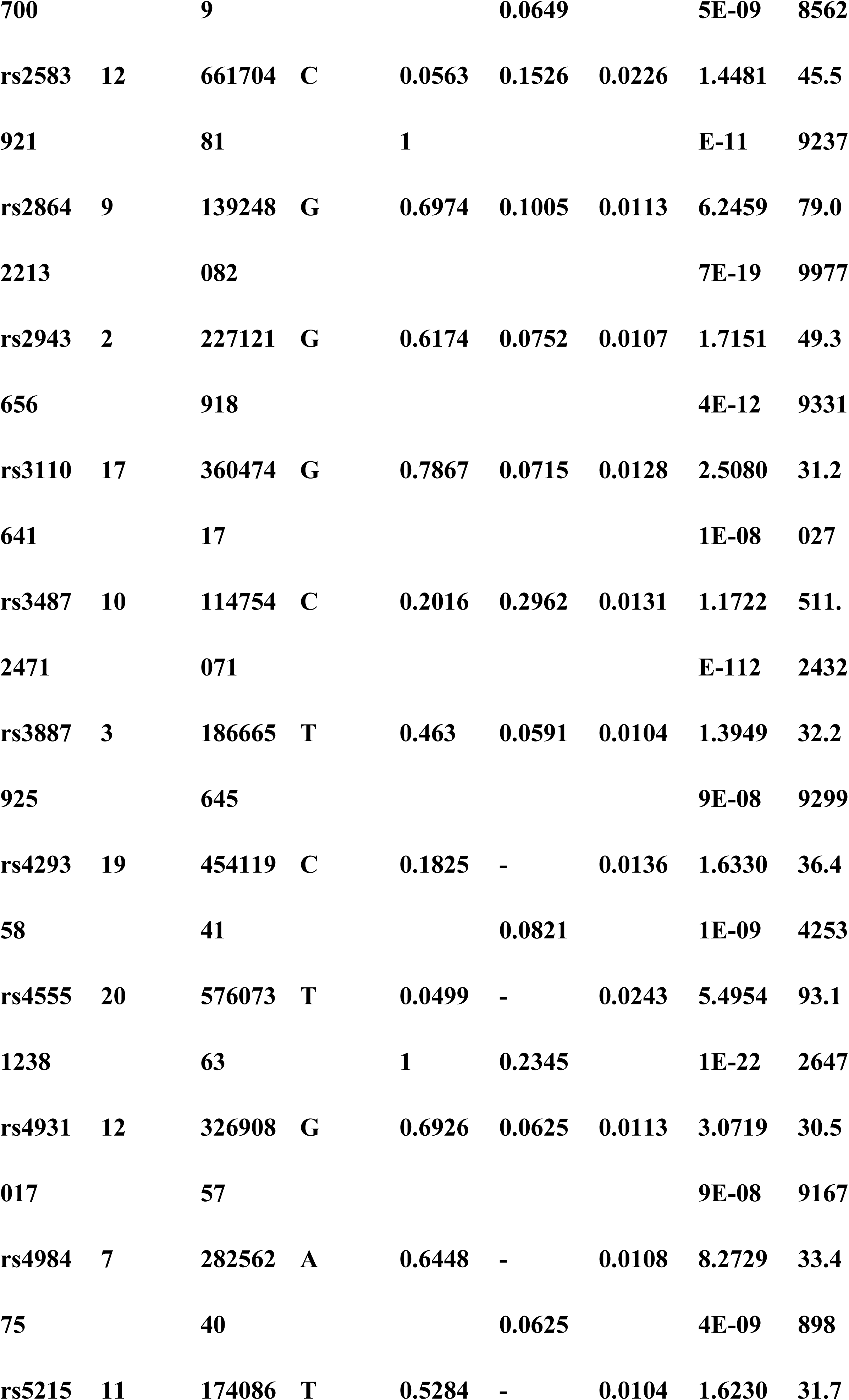
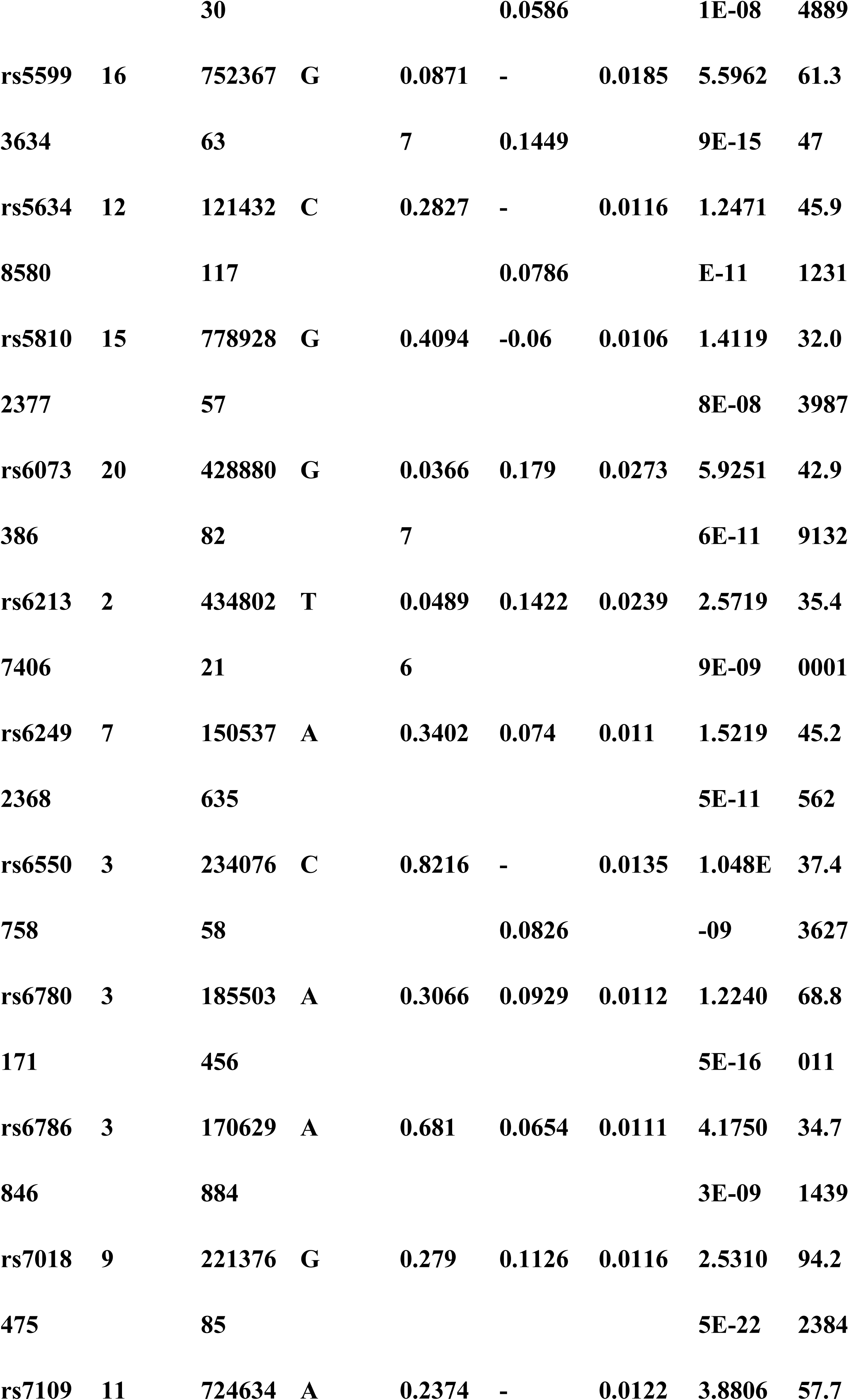
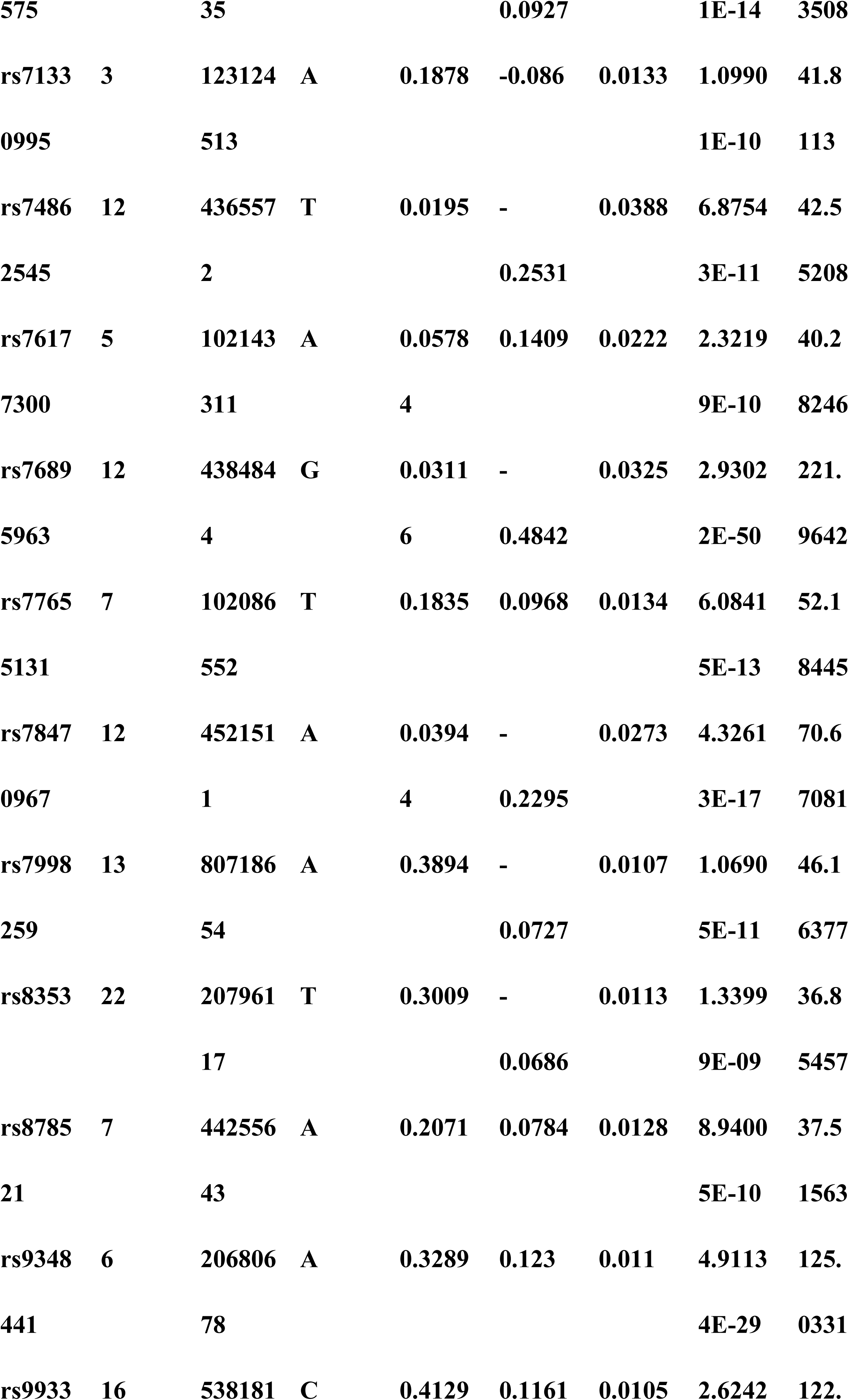
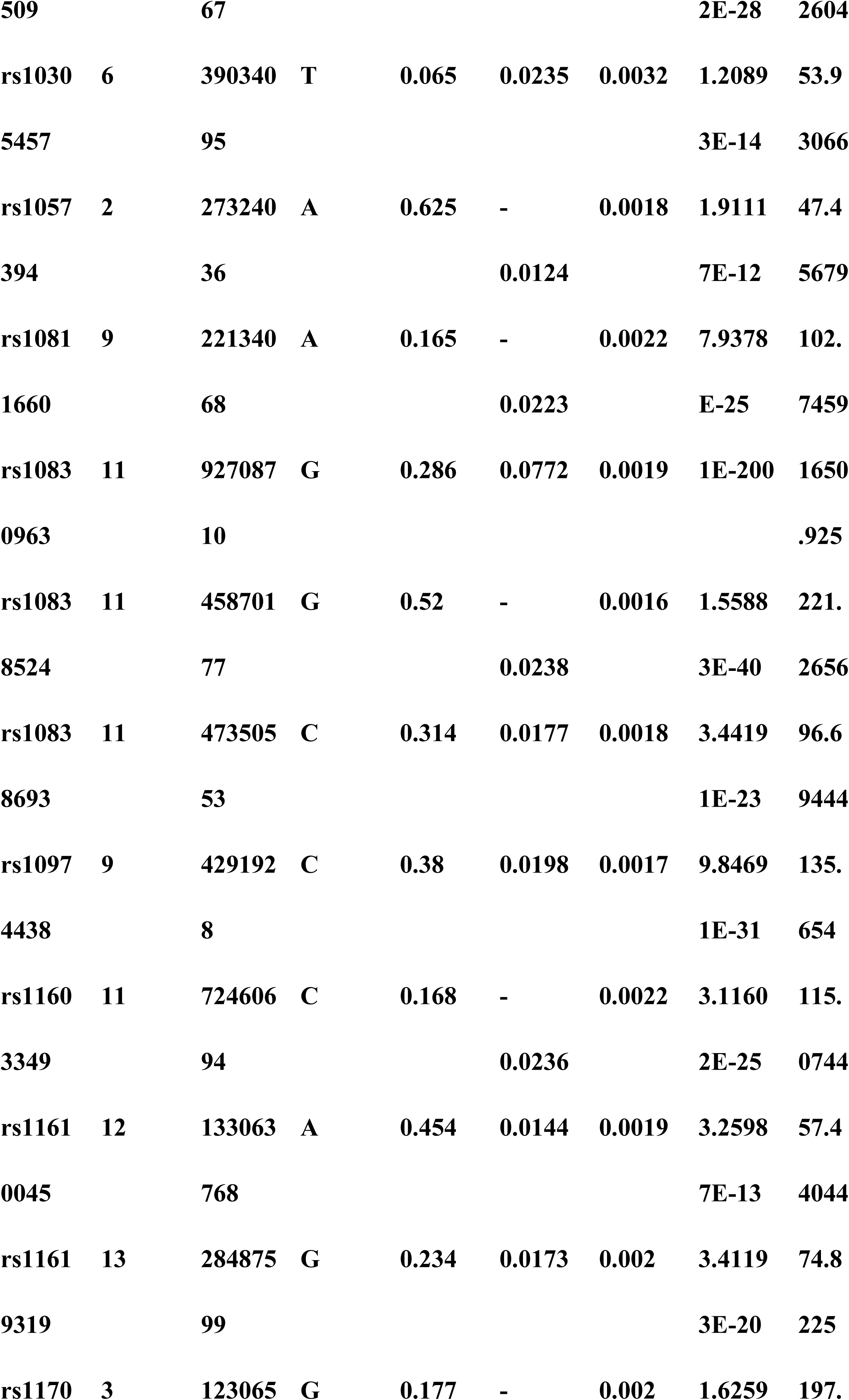
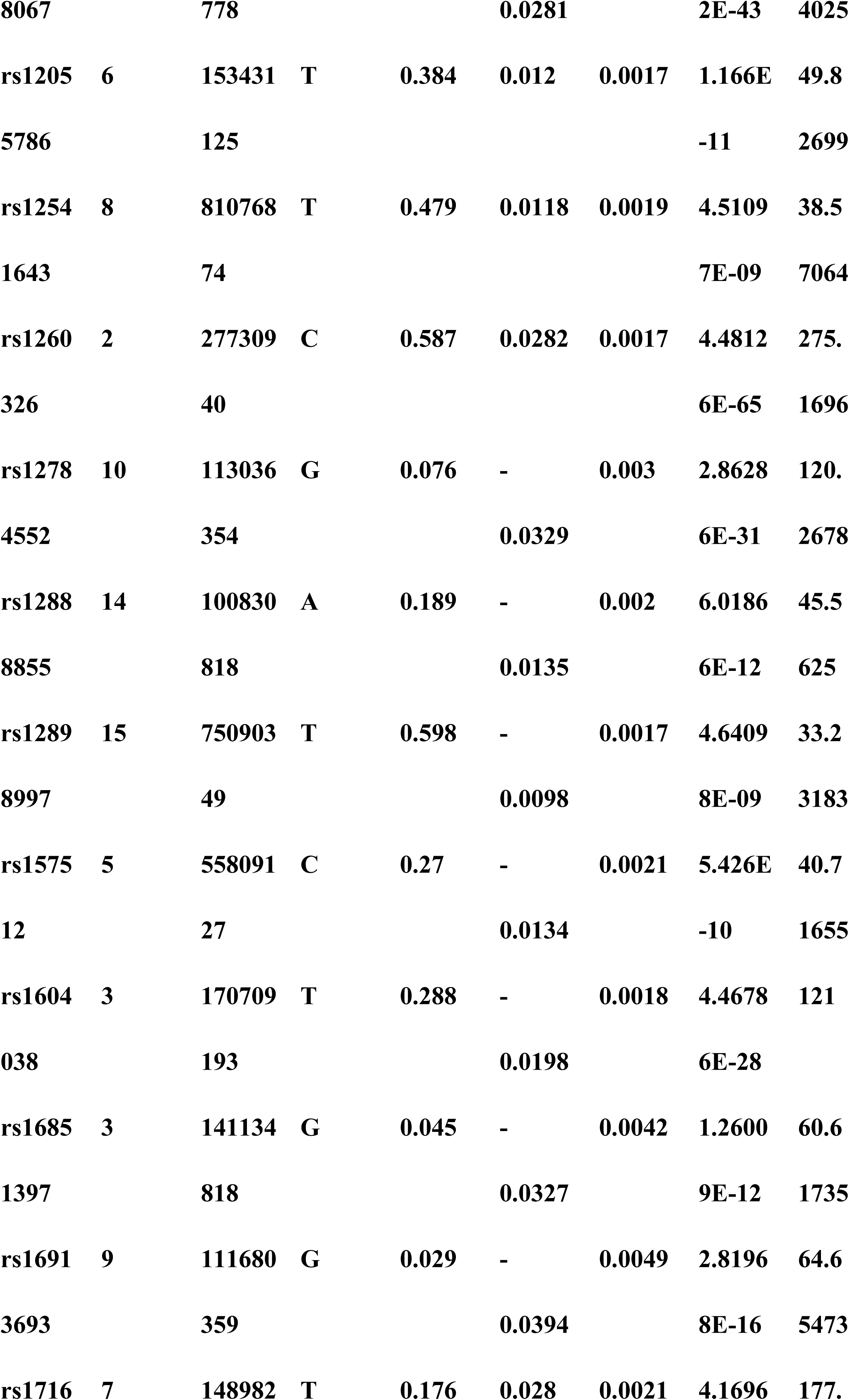
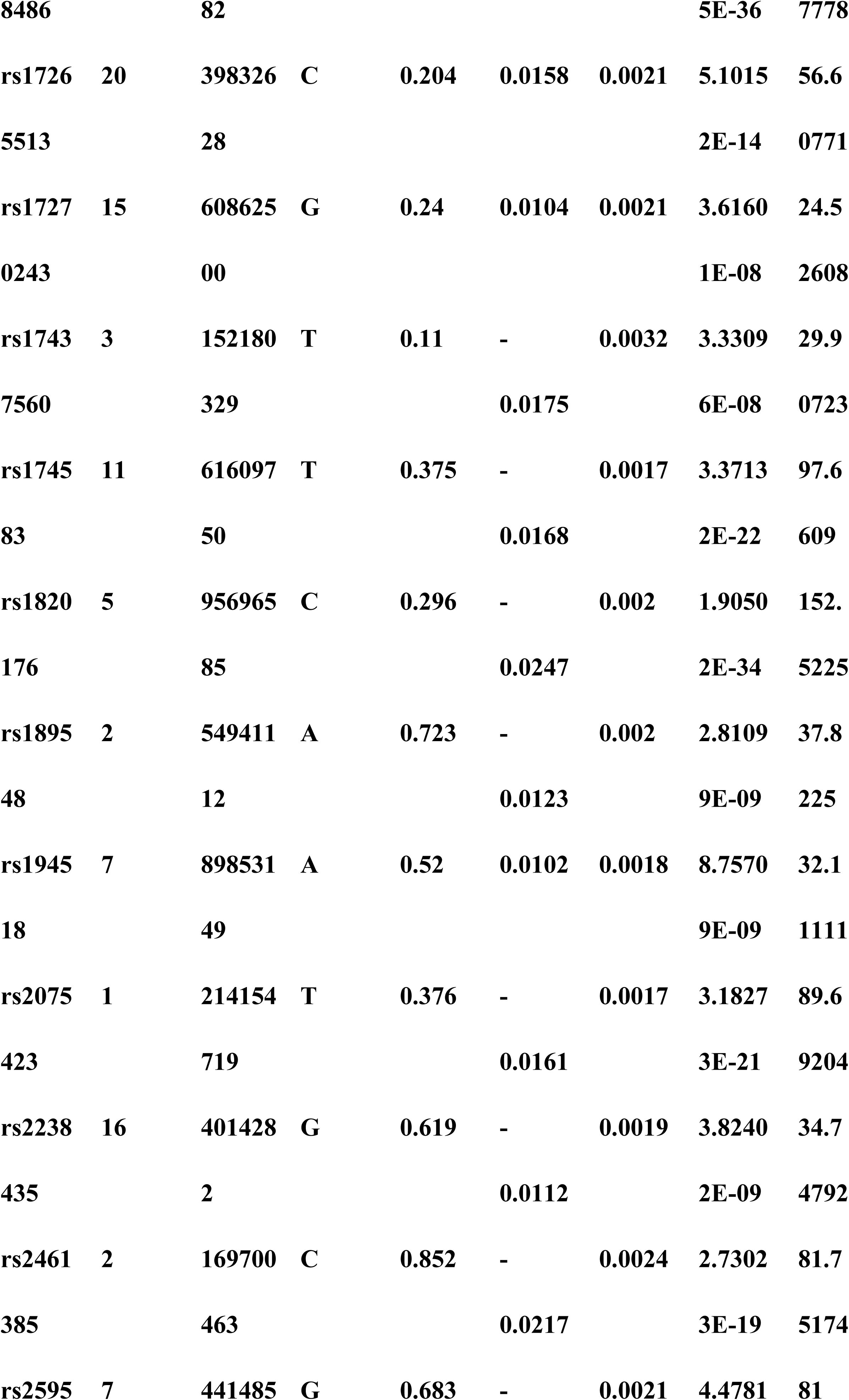
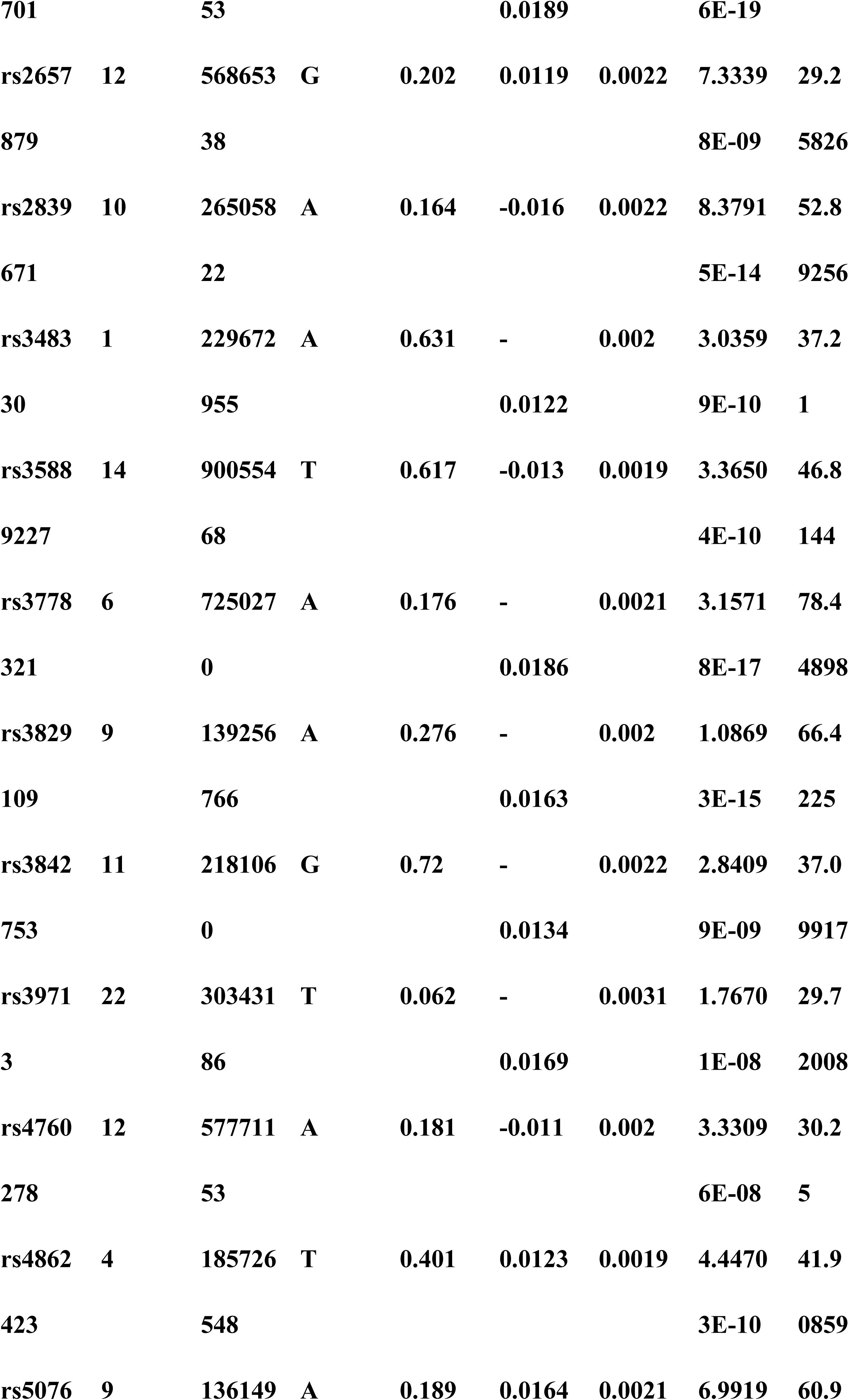
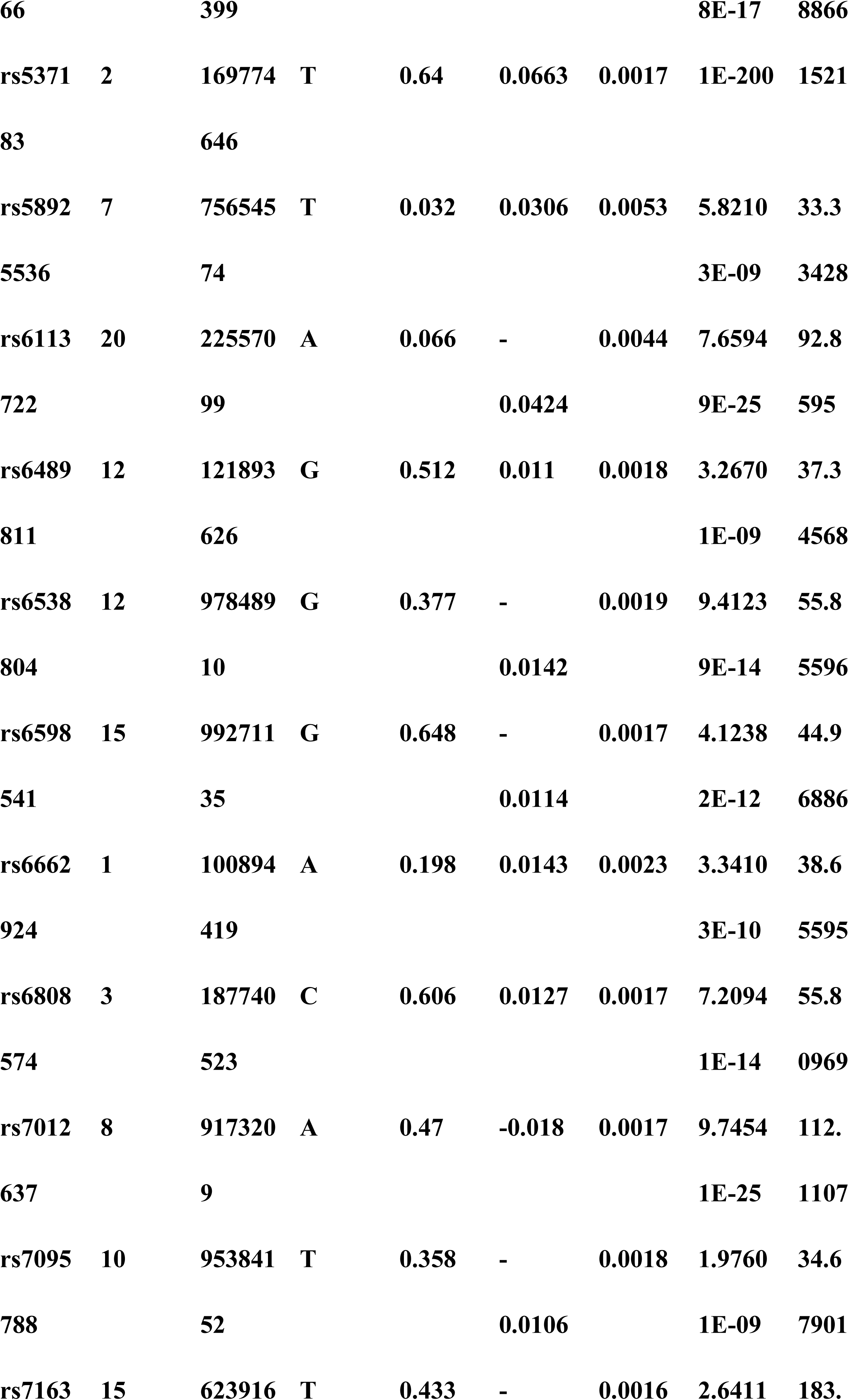
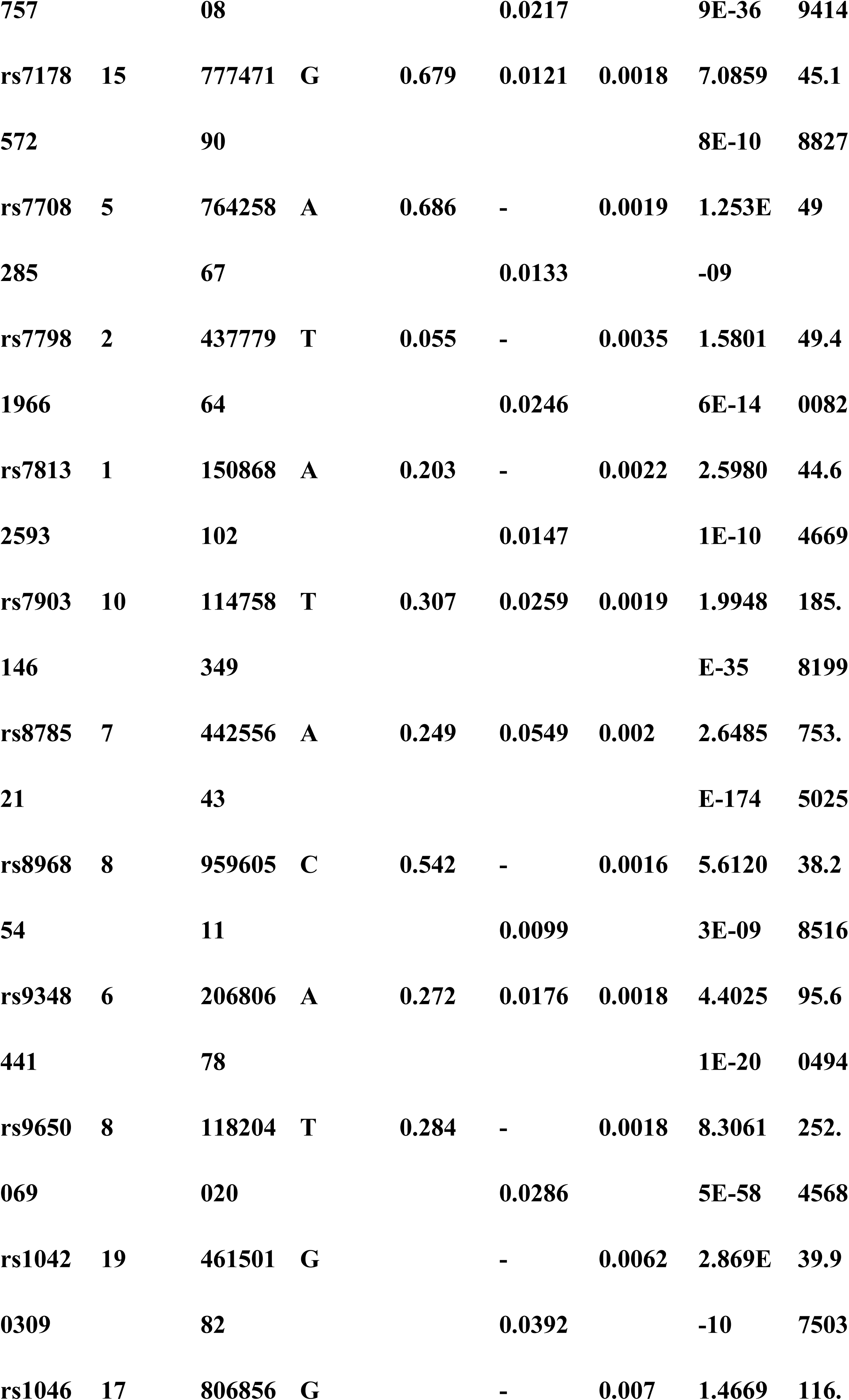
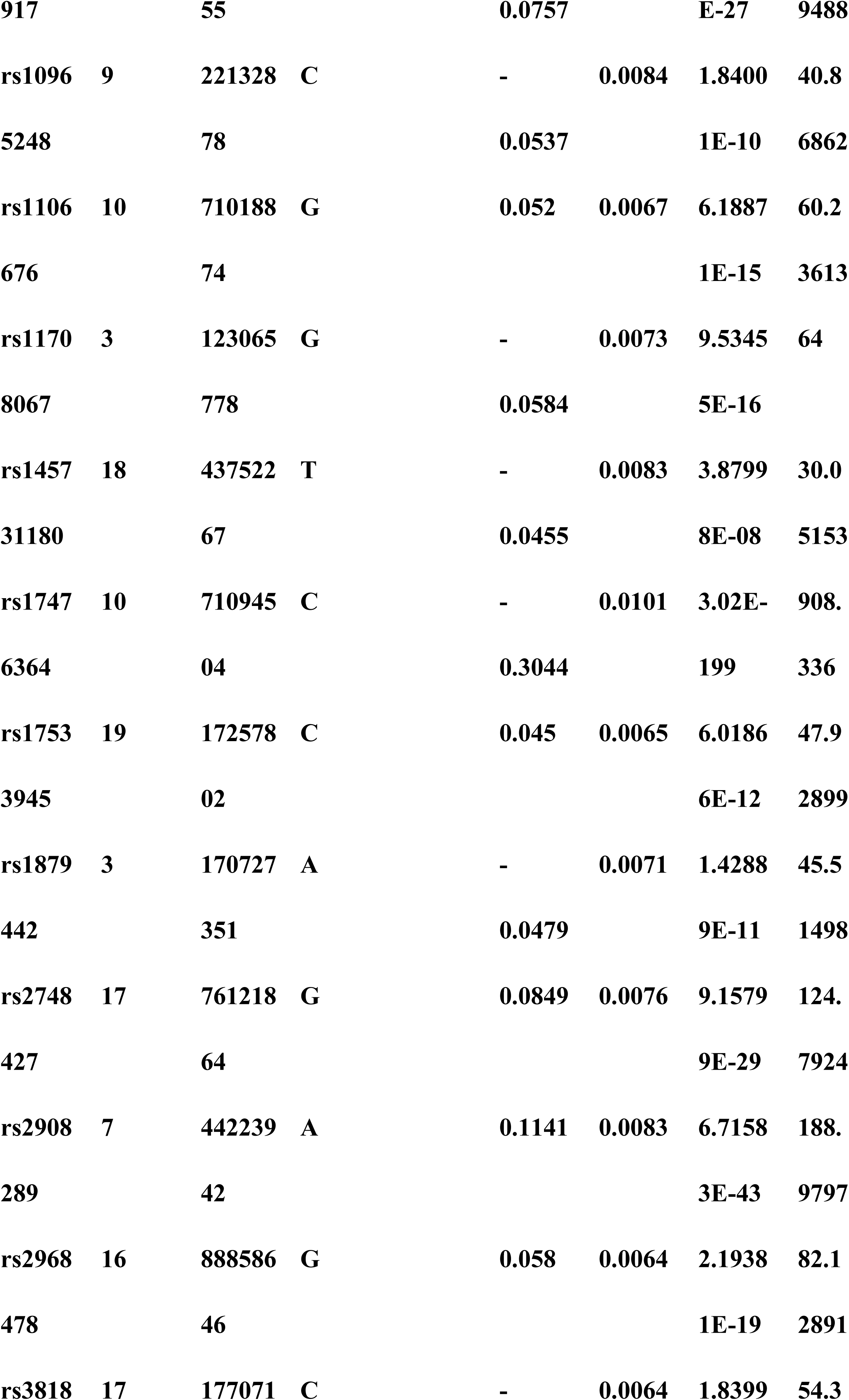
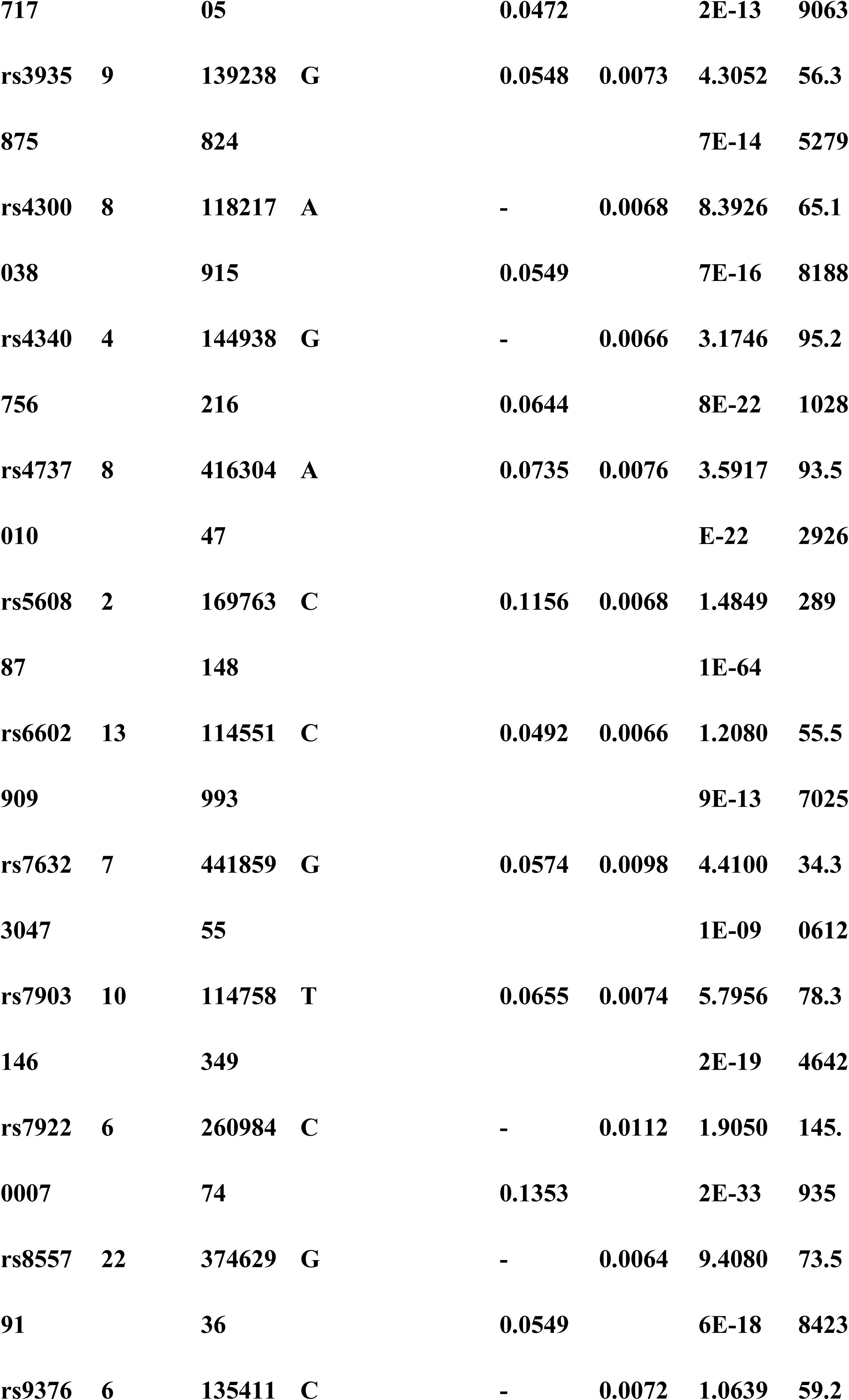
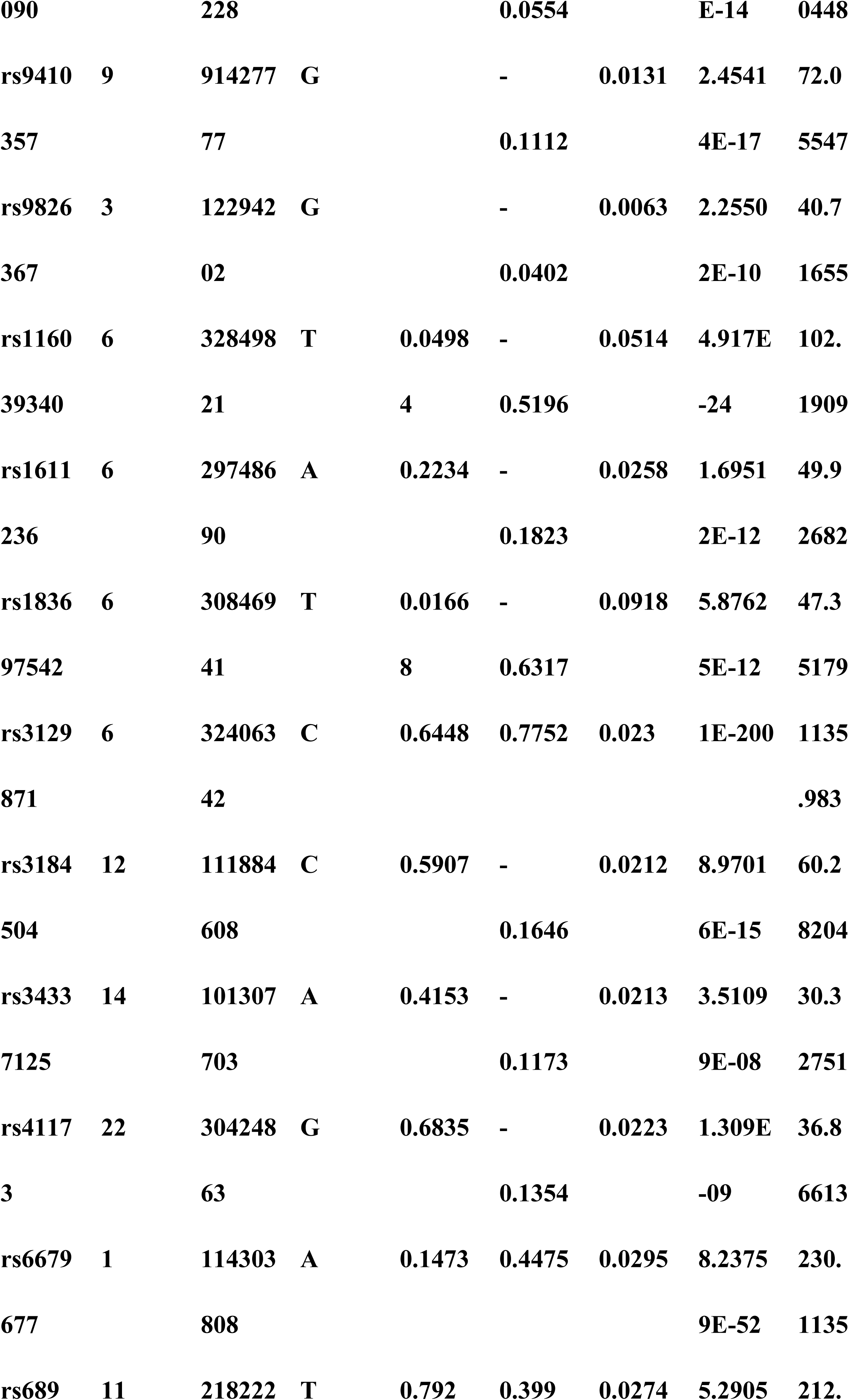
Genetic variants that were used as instruments for diabetes traits and blood sugar traits.

**Supplementary Table 3.** Genetic variants that were used as instruments for vilitigo.

| SNP | C<br>hr | Pos | Effe<br>ct<br>Alle<br>le | Effect<br>Allele<br>Freque<br>ncy | Beta | SE | P<br>valu<br>e | F |
| --- | --- | --- | --- | --- | --- | --- | --- | --- |
| rs108184 | 9 | 123634 | G | 0.01824 | 0.000860 | 0.000154 | 2.67 | 30.935 |
| 77 |  | 029 |  | 44 | 426 | 697 | E-08 | 936 |
| rs113405 | 7 | 644931 | T | 0.00325 | 0.002188 | 0.000383 | 1.15 | 32.577 |
| 464 |  |  |  | 031 | 17 | 374 | E-08 | 429 |
| rs114423 | 2 | 457693 | A | 0.00328 | 0.002186 | 0.000380 | 9.11 | 33.023 |
| 859 |  | 59 |  | 441 | 11 | 416 | E-09 | 756 |
| rs115241 | 2 | 203383 | C | 0.00856 | 0.001424 | 0.000228 | 4.91 | 38.716 |
| 301 |  | 263 |  | 171 | 19 | 886 | E-10 | 617 |
| rs117217<br>780 | 20 | 142924<br>67 | G | 0.01397<br>25 | 0.001036<br>9 | 0.000179<br>305 | 7.35<br>E-09 | 33.441<br>746 |
| rs117918<br>443 | 15 | 550961<br>57 | C | 0.00983<br>544 | 0.001390<br>76 | 0.000209<br>726 | 3.33<br>E-11 | 43.974<br>391 |
| rs132701<br>91 | 8 | 413325<br>11 | G | 0.01730<br>73 | 0.001112<br>81 | 0.000160<br>579 | 4.22<br>E-12 | 48.024<br>687 |
| rs138415<br>604 | 2 | 134190<br>944 | T | 0.02434<br>47 | 0.000769<br>211 | 0.000138<br>56 | 2.83<br>E-08 | 30.818<br>764 |
| rs140098<br>080 | 1 | 447721<br>00 | A | 0.00902<br>138 | 0.001236<br>13 | 0.000221<br>553 | 2.42<br>E-08 | 31.129<br>563 |
| rs141164<br>669 | 9 | 875216<br>26 | A | 0.00774<br>024 | 0.001552<br>91 | 0.000244<br>765 | 2.23<br>E-10 | 40.252<br>6 |
| rs144017<br>585 | 9 | 898405<br>75 | T | 0.00975<br>537 | 0.001312<br>13 | 0.000212<br>73 | 6.92<br>E-10 | 38.044<br>883 |
| rs145651<br>908 | 8 | 135649<br>198 | A | 0.00318<br>803 | 0.002205<br>27 | 0.000378<br>826 | 5.84<br>E-09 | 33.887<br>847 |
| rs147122<br>750 | 5 | 234518<br>05 | G | 0.01637<br>76 | 0.000937<br>204 | 0.000167<br>643 | 2.27<br>E-08 | 31.253<br>411 |
| rs147843<br>757 | 10 | 943823<br>59 | A | 0.02850<br>25 | 0.000686<br>841 | 0.000125<br>004 | 3.92<br>E-08 | 30.190<br>104 |
| rs148393 | 10 | 143321 | A | 0.02631 | 0.000751 | 0.000130 | 8.43 | 33.175 |
| 137 |  | 99 |  | 68 | 015 | 389 | E-09 | 335 |
| rs149961 | 19 | 542695 | C | 0.00608 | 0.001581 | 0.000279 | 1.52 | 32.028 |
| 991 |  | 12 |  | 543 | 49 | 446 | E-08 | 537 |
| rs151035 | 2 | 161261 | A | 0.00711 | 0.001438 | 0.000249 | 8.48 | 33.164 |
| 772 |  | 546 |  | 746 | 69 | 822 | E-09 | 472 |
| rs169420 | 18 | 235833 | G | 0.00454 | 0.001684 | 0.000304 | 3.14 | 30.621 |
| 56 |  | 15 |  | 183 | 31 | 376 | E-08 | 273 |
| rs181638 | 4 | 404169 | A | 0.00606 | 0.001486 | 0.000272 | 4.71 | 29.835 |
| 428 |  | 41 |  | 912 | 13 | 076 | E-08 | 487 |
| rs482381 | 22 | 468350 | C | 0.00100 | 0.004158 | 0.000645 | 1.19 | 41.483 |
| 0 |  | 00 |  | 089 | 63 | 674 | E-10 | 364 |
| rs765297 | 9 | 757277 | G | 0.00709 | 0.001607 | 0.000243 | 4.29 | 43.480 |
| 09 |  | 48 |  | 225 | 31 | 754 | E-11 | 626 |
| rs108184 | 9 | 123634 | G | 0.01824 | 0.000860 | 0.000154 | 2.67 | 30.935 |
| 77 |  | 029 |  | 44 | 426 | 697 | E-08 | 936 |
| rs113405 | 7 | 644931 | T | 0.00325 | 0.002188 | 0.000383 | 1.15 | 32.577 |
| 464 |  |  |  | 031 | 17 | 374 | E-08 | 429 |
| rs114423 | 2 | 457693 | A | 0.00328 | 0.002186 | 0.000380 | 9.11 | 33.023 |
| 859 |  | 59 |  | 441 | 11 | 416 | E-09 | 756 |
| rs115241 | 2 | 203383 | C | 0.00856 | 0.001424 | 0.000228 | 4.91 | 38.716 |
| 301 |  | 263 |  | 171 | 19 | 886 | E-10 | 617 |
| rs117217<br>780 | 20 | 142924<br>67 | G | 0.01397<br>25 | 0.001036<br>9 | 0.000179<br>305 | 7.35<br>E-09 | 33.441<br>746 |
| rs117918<br>443 | 15 | 550961<br>57 | C | 0.00983<br>544 | 0.001390<br>76 | 0.000209<br>726 | 3.33<br>E-11 | 43.974<br>391 |
| rs132701<br>91 | 8 | 413325<br>11 | G | 0.01730<br>73 | 0.001112<br>81 | 0.000160<br>579 | 4.22<br>E-12 | 48.024<br>687 |
| rs138415<br>604 | 2 | 134190<br>944 | T | 0.02434<br>47 | 0.000769<br>211 | 0.000138<br>56 | 2.83<br>E-08 | 30.818<br>764 |
| rs140098<br>080 | 1 | 447721<br>00 | A | 0.00902<br>138 | 0.001236<br>13 | 0.000221<br>553 | 2.42<br>E-08 | 31.129<br>563 |
| rs141164<br>669 | 9 | 875216<br>26 | A | 0.00774<br>024 | 0.001552<br>91 | 0.000244<br>765 | 2.23<br>E-10 | 40.252<br>6 |
| rs144017<br>585 | 9 | 898405<br>75 | T | 0.00975<br>537 | 0.001312<br>13 | 0.000212<br>73 | 6.92<br>E-10 | 38.044<br>883 |
| rs147122<br>750 | 5 | 234518<br>05 | G | 0.01637<br>76 | 0.000937<br>204 | 0.000167<br>643 | 2.27<br>E-08 | 31.253<br>411 |
| rs147843<br>757 | 10 | 943823<br>59 | A | 0.02850<br>25 | 0.000686<br>841 | 0.000125<br>004 | 3.92<br>E-08 | 30.190<br>104 |
| rs148393<br>137 | 10 | 143321<br>99 | A | 0.02631<br>68 | 0.000751<br>015 | 0.000130<br>389 | 8.43<br>E-09 | 33.175<br>335 |
| rs149961 | 19 | 542695 | C | 0.00608 | 0.001581 | 0.000279 | 1.52 | 32.028 |
| 991 |  | 12 |  | 543 | 49 | 446 | E-08 | 537 |
| rs151035 | 2 | 161261 | A | 0.00711 | 0.001438 | 0.000249 | 8.48 | 33.164 |
| 772 |  | 546 |  | 746 | 69 | 822 | E-09 | 472 |
| rs169420 | 18 | 235833 | G | 0.00454 | 0.001684 | 0.000304 | 3.14 | 30.621 |
| 56 |  | 15 |  | 183 | 31 | 376 | E-08 | 273 |
| rs181638 | 4 | 404169 | A | 0.00606 | 0.001486 | 0.000272 | 4.71 | 29.835 |
| 428 |  | 41 |  | 912 | 13 | 076 | E-08 | 487 |
| rs482381 | 22 | 468350 | C | 0.00100 | 0.004158 | 0.000645 | 1.19 | 41.483 |
| 0 |  | 00 |  | 089 | 63 | 674 | E-10 | 364 |
| rs765297 | 9 | 757277 | G | 0.00709 | 0.001607 | 0.000243 | 4.29 | 43.480 |
| 09 |  | 48 |  | 225 | 31 | 754 | E-11 | 626 |
| rs108184 | 9 | 123634 | G | 0.01824 | 0.000860 | 0.000154 | 2.67 | 30.935 |
| 77 |  | 029 |  | 44 | 426 | 697 | E-08 | 936 |
| rs113405 | 7 | 644931 | T | 0.00325 | 0.002188 | 0.000383 | 1.15 | 32.577 |
| 464 |  |  |  | 031 | 17 | 374 | E-08 | 429 |
| rs114423 | 2 | 457693 | A | 0.00328 | 0.002186 | 0.000380 | 9.11 | 33.023 |
| 859 |  | 59 |  | 441 | 11 | 416 | E-09 | 756 |
| rs115241 | 2 | 203383 | C | 0.00856 | 0.001424 | 0.000228 | 4.91 | 38.716 |
| 301 |  | 263 |  | 171 | 19 | 886 | E-10 | 617 |
| rs117217 | 20 | 142924 | G | 0.01397 | 0.001036 | 0.000179 | 7.35 | 33.441 |
| 780 |  | 67 |  | 25 | 9 | 305 | E-09 | 746 |
| rs117918<br>443 | 15 | 550961<br>57 | C | 0.00983<br>544 | 0.001390<br>76 | 0.000209<br>726 | 3.33<br>E-11 | 43.974<br>391 |
| rs132701<br>91 | 8 | 413325<br>11 | G | 0.01730<br>73 | 0.001112<br>81 | 0.000160<br>579 | 4.22<br>E-12 | 48.024<br>687 |
| rs138415<br>604 | 2 | 134190<br>944 | T | 0.02434<br>47 | 0.000769<br>211 | 0.000138<br>56 | 2.83<br>E-08 | 30.818<br>764 |
| rs140098<br>080 | 1 | 447721<br>00 | A | 0.00902<br>138 | 0.001236<br>13 | 0.000221<br>553 | 2.42<br>E-08 | 31.129<br>563 |
| rs141164<br>669 | 9 | 875216<br>26 | A | 0.00774<br>024 | 0.001552<br>91 | 0.000244<br>765 | 2.23<br>E-10 | 40.252<br>6 |
| rs144017<br>585 | 9 | 898405<br>75 | T | 0.00975<br>537 | 0.001312<br>13 | 0.000212<br>73 | 6.92<br>E-10 | 38.044<br>883 |
| rs147122<br>750 | 5 | 234518<br>05 | G | 0.01637<br>76 | 0.000937<br>204 | 0.000167<br>643 | 2.27<br>E-08 | 31.253<br>411 |
| rs147843<br>757 | 10 | 943823<br>59 | A | 0.02850<br>25 | 0.000686<br>841 | 0.000125<br>004 | 3.92<br>E-08 | 30.190<br>104 |
| rs148393<br>137 | 10 | 143321<br>99 | A | 0.02631<br>68 | 0.000751<br>015 | 0.000130<br>389 | 8.43<br>E-09 | 33.175<br>335 |
| rs149961<br>991 | 19 | 542695<br>12 | C | 0.00608<br>543 | 0.001581<br>49 | 0.000279<br>446 | 1.52<br>E-08 | 32.028<br>537 |
| rs151035 | 2 | 161261 | A | 0.00711 | 0.001438 | 0.000249 | 8.48 | 33.164 |
| 772 |  | 546 |  | 746 | 69 | 822 | E-09 | 472 |
| rs169420 | 18 | 235833 | G | 0.00454 | 0.001684 | 0.000304 | 3.14 | 30.621 |
| 56 |  | 15 |  | 183 | 31 | 376 | E-08 | 273 |
| rs181638 | 4 | 404169 | A | 0.00606 | 0.001486 | 0.000272 | 4.71 | 29.835 |
| 428 |  | 41 |  | 912 | 13 | 076 | E-08 | 487 |
| rs482381 | 22 | 468350 | C | 0.00100 | 0.004158 | 0.000645 | 1.19 | 41.483 |
| 0 |  | 00 |  | 089 | 63 | 674 | E-10 | 364 |
| rs765297 | 9 | 757277 | G | 0.00709 | 0.001607 | 0.000243 | 4.29 | 43.480 |
| 09 |  | 48 |  | 225 | 31 | 754 | E-11 | 626 |
| rs108184 | 9 | 123634 | G | 0.01824 | 0.000860 | 0.000154 | 2.67 | 30.935 |
| 77 |  | 029 |  | 44 | 426 | 697 | E-08 | 936 |
| rs117217 | 20 | 142924 | G | 0.01397 | 0.001036 | 0.000179 | 7.35 | 33.441 |
| 780 |  | 67 |  | 25 | 9 | 305 | E-09 | 746 |
| rs132701 | 8 | 413325 | G | 0.01730 | 0.001112 | 0.000160 | 4.22 | 48.024 |
| 91 |  | 11 |  | 73 | 81 | 579 | E-12 | 687 |
| rs138415 | 2 | 134190 | T | 0.02434 | 0.000769 | 0.000138 | 2.83 | 30.818 |
| 604 |  | 944 |  | 47 | 211 | 56 | E-08 | 764 |
| rs147122 | 5 | 234518 | G | 0.01637 | 0.000937 | 0.000167 | 2.27 | 31.253 |
| 750 |  | 05 |  | 76 | 204 | 643 | E-08 | 411 |
| rs147843 | 10 | 943823 | A | 0.02850 | 0.000686 | 0.000125 | 3.92 | 30.190 |
| 757 |  | 59 |  | 25 | 841 | 004 | E-08 | 104 |
| rs148393 | 10 | 143321 | A | 0.02631 | 0.000751 | 0.000130 | 8.43 | 33.175 |
| 137 |  | 99 |  | 68 | 015 | 389 | E-09 | 335 |

### Univariate MR Analysis

Glycemic traits (fasting blood glucose; Hba1c), diabetes traits (T1DM; T2DM) and vitiligo. IVW method was used as the main test method, and the results showed as S3Table; Cochran’s Q(18) was used as the heterogeneity test, and the results showed that the Qpval value was < 0.05, suggesting that there was no heterogeneity. Therefore, we used the fixed effects model IVW approach for the study. The specific results are shown in S4 table. Mendelian randomisation results of causal effects between blood glucose characteristics, diabetes mellitus and vitiligo.The figure show that T1DM has a causal relationship with the onset of vitiligo (p=0.018; 95%OR:1.000(1.000-1.000)). In sensitivity analysis (MR- Egger,Weighted Mode,Weighted Median), horizontal diversity was not found (p=0.018; 95%OR: 1.000-1.000). We performed Leaveone analysis on the results to evaluate the influence of single SNPs on the outcome, and the results are shown in S2Fig.

**Supplementary Table 4.** Unltivariate mendelian randomisation results of causal efects between blood glucose characteristics, diabetes mellitus and vitiligo (p < 5 × 10−8)

| outcome | exposure | method | nsnp | pval |
| --- | --- | --- | --- | --- |
| vitiligo | T1DM | Inverse variance weighted | 13 | 0.018 |
|  |  | MR Egger |  | 0.625 |
|  |  | Weighted median |  | 0.543 |
|  |  | Weighted mode |  | 0.231 |
|  | T2DM | Inverse variance weighted | 56 | 0.878 |
|  |  | MR Egger |  | 0.914 |
|  |  | Weighted median |  | 0.61 |
|  |  | Weighted mode |  | 0.712 |
|  | Fasting glucose | Inverse variance weighted | 64 | 0.247 |
|  |  | MR Egger |  | 0.269 |
|  |  | Weighted median |  | 0.40 |
|  |  | Weighted mode |  | 0.326 |
|  | HbA1c | Inverse variance weighted | 26 | 0.303 |
|  |  | MR Egger |  | 0.313 |
|  |  | Weighted median |  | 0.107 |
|  |  | Weighted mode |  | 0.18 |

### Multivariate MR Analysis

Based on the single-sample MR Analysis, we constructed a multivariate MR Model to analyze the causal relationship between Hba1c and T1DM and vitiligo. Associations were assessed using the multivariate IVW method as the primary analysis method. The results showed that there was still a significant causal association between T1DM and the onset of vitiligo (p=0.016, 95% OR= 1.000(1.000-1.000)), which was consistent with the results of univariate MR Analysis(S5Table).Horizontal pleiotropy was tested by MR-PRESSO method, and the results were consistent with the IVW results, and there was no horizontal pleiotropy.

**Supplementary Table 5.**
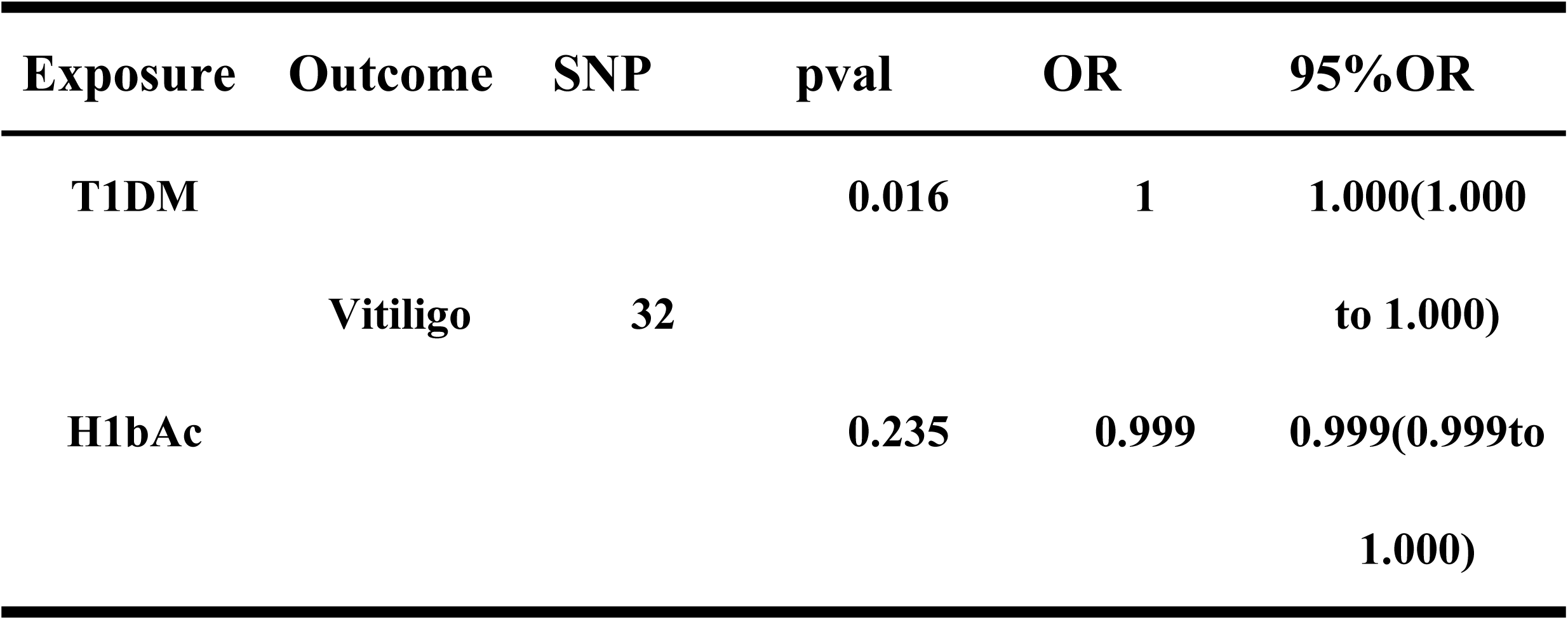
Multivariate mendelian randomisation results of causal efects between Type 1 diabetes, glycated haemoglobin and vitiligo (p < 5 × 10−8)

### Reverse MR Analysis

After the univariate MR Analysis demonstrated the causal association of T1DM in the pathogenesis of vitiligo, we further explored the causal association between vitiligo and blood glucose features and diabetes features. To this end, we performed reverse MR Analysis, the specific methods and procedures were consistent with univariate MR Analysis. The results of IVW method are shown in S10, with vitiligo as exposure, blood glucose features and glycosuria There were no significant causal associations between disease characteristics. S3 Fig.

**Table S6.**
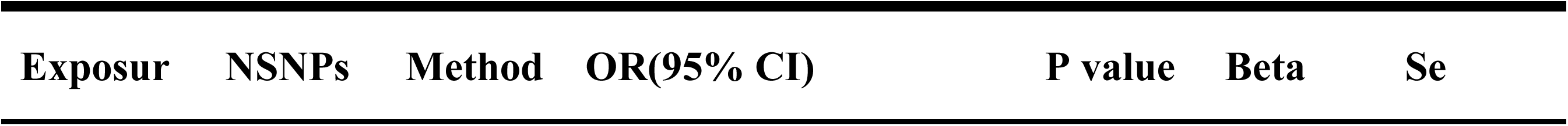
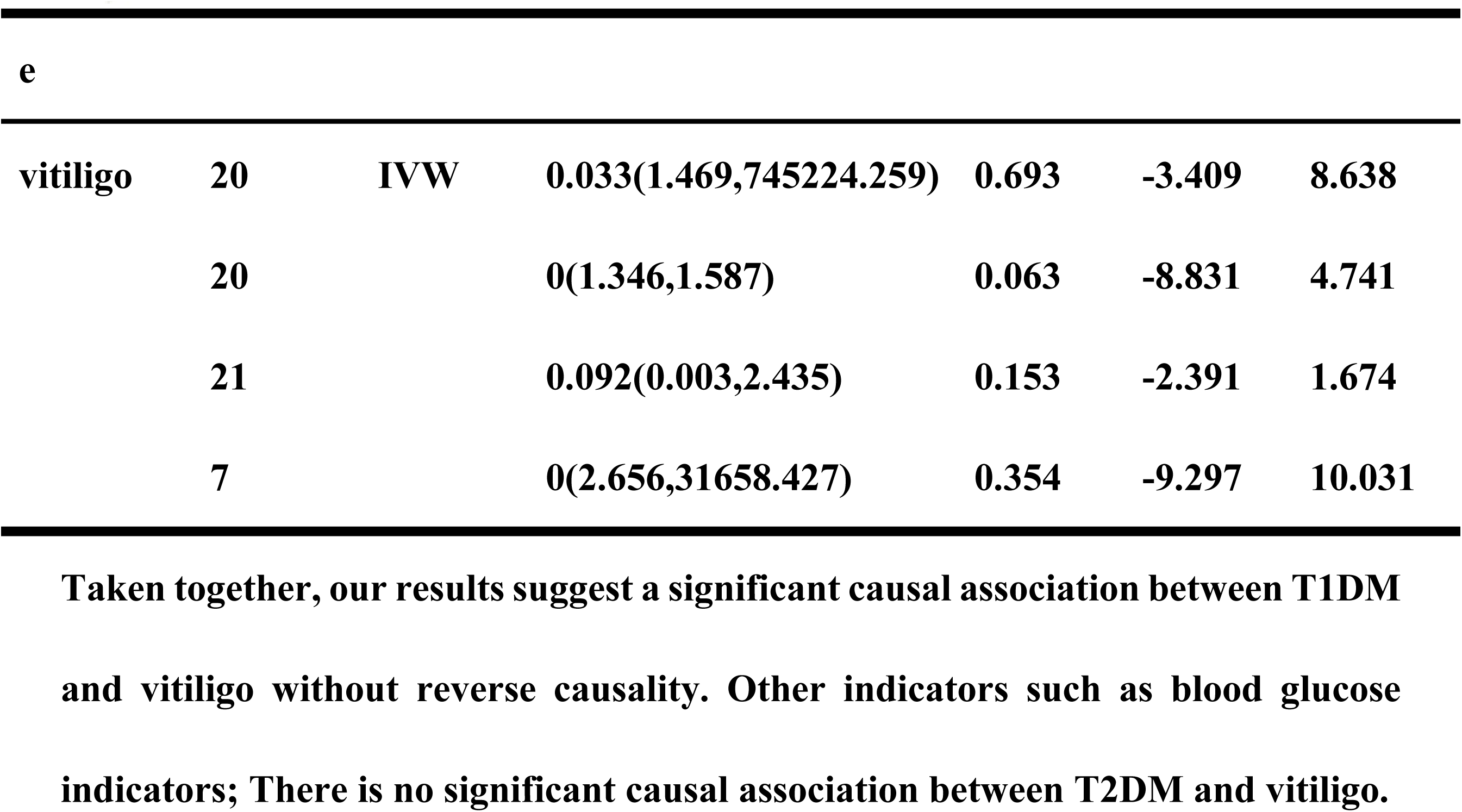
Reverse mendelian randomisation results of causal efects between vitiligo and Type 1 diabetes,Type 2 diabetes, glycated haemoglobin, fasting blood glucose (p < 5 × 10−8)

| Exposur | NSNPs | Method | OR(95% CI) | P value | Beta | Se |
| --- | --- | --- | --- | --- | --- | --- |
| e |  |  |  |  |  |  |
| vitiligo | 20 | IVW | 0.033(1.469,745224.259) | 0.693 | -3.409 | 8.638 |
|  | 20 |  | 0(1.346,1.587) | 0.063 | -8.831 | 4.741 |
|  | 21 |  | 0.092(0.003,2.435) | 0.153 | -2.391 | 1.674 |
|  | 7 |  | 0(2.656,31658.427) | 0.354 | -9.297 | 10.031 |

### Reverse MR leave-one-out sensitivity analysis

Taken together, our results suggest a significant causal association between T1DM and vitiligo without reverse causality. Other indicators such as blood glucose indicators; There is no significant causal association between T2DM and vitiligo.

## Discussion

Based on the results of this experiment, we conclude that there is a positive causal association between diabetes and the occurrence of vitiligo in T1 stage, and there is no negative causal association between diabetes and blood glucose traits in T2 stage and the occurrence of vitiligo.

### Assumptions to explain the outcomes

The method used in this study is bidirectional MR Analysis(19) combined with TSMR analysis, which has the advantage of further avoiding the interference of horizontal pleiotropy and heterogeneity on the study results compared with single MR Analysis. Pleiotropy refers to genetic variants as SNPs that may be associated with multiple phenotypes, which can lead to biased causal predictions from MR Analysis.

A number of studies(20) have shown that diabetes induces different degrees of metabolic problems, and many confounding factors may affect the outcome. Some observational studies(4) have suggested that T1DM and T2DM are closely related to the development of vitiligo. However, in our study, our results do not support an association between T2DM and the onset of vitiligo. Because of the complex pathogenesis of diabetes and its tendency to be complicated by other disease complications, we believe that the association between type T2DM and vitiligo mentioned in observational studies may be related to confounding factors. Two MR Analyses(21, 22) suggested a significant causal association between T2DM and depression, and one meta-analysis(23) concluded that vitiligo induces varying degrees of psychological problems. This finding may be an important reason why vitiligo is associated with T2DM in several observational studies.

To investigate the relationship between blood glucose and diabetes characteristics and the pathogenesis of vitiligo. Supported by the available data, we conclude that T1DM is causally related to vitiligo and that T1DM does not affect the onset of vitiligo using Hba1c as a mediator.

Type 1 diabetes mellitus (T1DM)(24) is an autoimmune disease that results in the destruction of insulin-producing pancreatic beta cells. Some studies suggest that T cells and B cells play a crucial role(25) in the pathogenesis of diseases. It has been shown(26) that β-cell-targeting autoantibodies targeting insulin or GAD65 can be detected in the serum early in the onset of type 1 diabetes. .In addition, CD4+ and CD8+ T cells that are specific for β-cell autoantigens are detectable in patients with stage 3 T1DM and even in patients with earlier stages of the disease(27, 28) . The specific differentiation of T cells may be one of the mechanisms of vitiligo induced by type 1 diabetes. Studies(29) have shown that CD8+ cells play an important role in the pathogenesis of vitiligo. On the one hand, production of the cytokine IFN-γ by CD8+ cells is central(29) to disease progression. T cells(30) secrete IFN-γ and induce keratinocytes to produce T cell chemokine receptor CXCR3 ligands such as CXCL9 and CXCL10, thereby increasing the number of T cells in the body and promoting the development of vitiligo. On the other hand, the concentration of melanocyte-specific CD8 cells(31) was significantly increased in the early histological examination of vitiligo lesions, and the clones generated by these cells could kill and pigmented cells in vitro(32). We therefore suggest that the proliferation of CD8T cells induced by T1DM may be a mediator of the effect of T1DM on the pathogenesis of vitiligo. We will explore this issue in a follow-up study.

Our findings are consistent with those of some controlled experiments(33). We provide theoretical support for this conclusion from the perspective of MR Analysis.

### Limitation analysis

The main potential limitations of this study are:

1. The analyzed population was limited to European populations, and since race may have influenced our results, we may need to conduct the same MR Study in other races for validation.

A controlled study(34) with 142 samples suggested an association between insulin resistance and vitiligo. We did not include data on insulin resistance in our GWAS database because the data were current for 2010 and the sample size was not large enough to support the MR Analysis. Since insulin resistance is one of the major clinical manifestations of type 2 diabetes, we believe that this may have had some influence on our study. Further studies on this trait will be conducted after the GWAS database updates the relevant data.

## Conclusion

The results of MR Studies have shown that T1DM is a potential risk factor for the development of vitiligo, and Mendelian randomization (MR) results have provided genetic evidence for a causal relationship between T1DM and vitiligo. Clinical use of measures to treat T1DM may be a new idea for the prevention or treatment of vitiligo.

## Author Contributions

Conceptualization, S.H.methodology, software, and investigation, Y.C,S.H.; validation,Y.C; formal analysis, S.H; data curation, Y.C. and S.H.; writing— original draft preparation, S.H; writing—review and editing, S.H., Y.C. and J.C.; visualization, Y.C; supervision, Y.C; project administration, J.G,J.Z.; All authors have read and agreed to the published version of the manuscript.

## Funding

This study was supported by the National Natural Science Foundation of China(grant nos. 82074443), and by the Program of Science and Technology Department of Sichuan Province (2021ZYD0089).

## Abbreviations and standard terms used in this study

MR: Mendelian randomization
T1DM: Type 1 Diabetes Mellitus
T2DM: Type 2 Diabetes Mellitus
SNPs: Single-nucleotide polymorphisms
NA: Not available
SE: standard error
OR: Odds ratio
CI: Confidence interval
IV: Instrumental variables
IVW: Inverse variance weighting
MR-PRESSO: Mendelian Randomization Pleiotropy RESidual Sum and Outlier.

## Acknowledgments

Special thanks to the IEU open GWAS project developed by The MRC Integrative Epidemiology Unit (IEU) at the University of Bristol.We want to acknowledge the participants and investigators of the FinnGen study.We would also like to acknowledge the summary statistics provided by the UK Biobank and the European Bioinformatics Institute.

## Data Availability Statement

Publicly available datasets were analyzed in this study. This data can be found here: All GWAS data used in this study are available in the IEU open GWAS project (https://gwas.mrcieu.ac.uk/). Detailed data on vitiligo are available in the GWAS through the UK Biobank data(http://www.nealelab.is/uk-biobank/c). Summary statistics for T1DM and T2DM are from the FinnGen Consortium (https://r8.finngen.fi).

## Supporting information

**S1 Fig.**
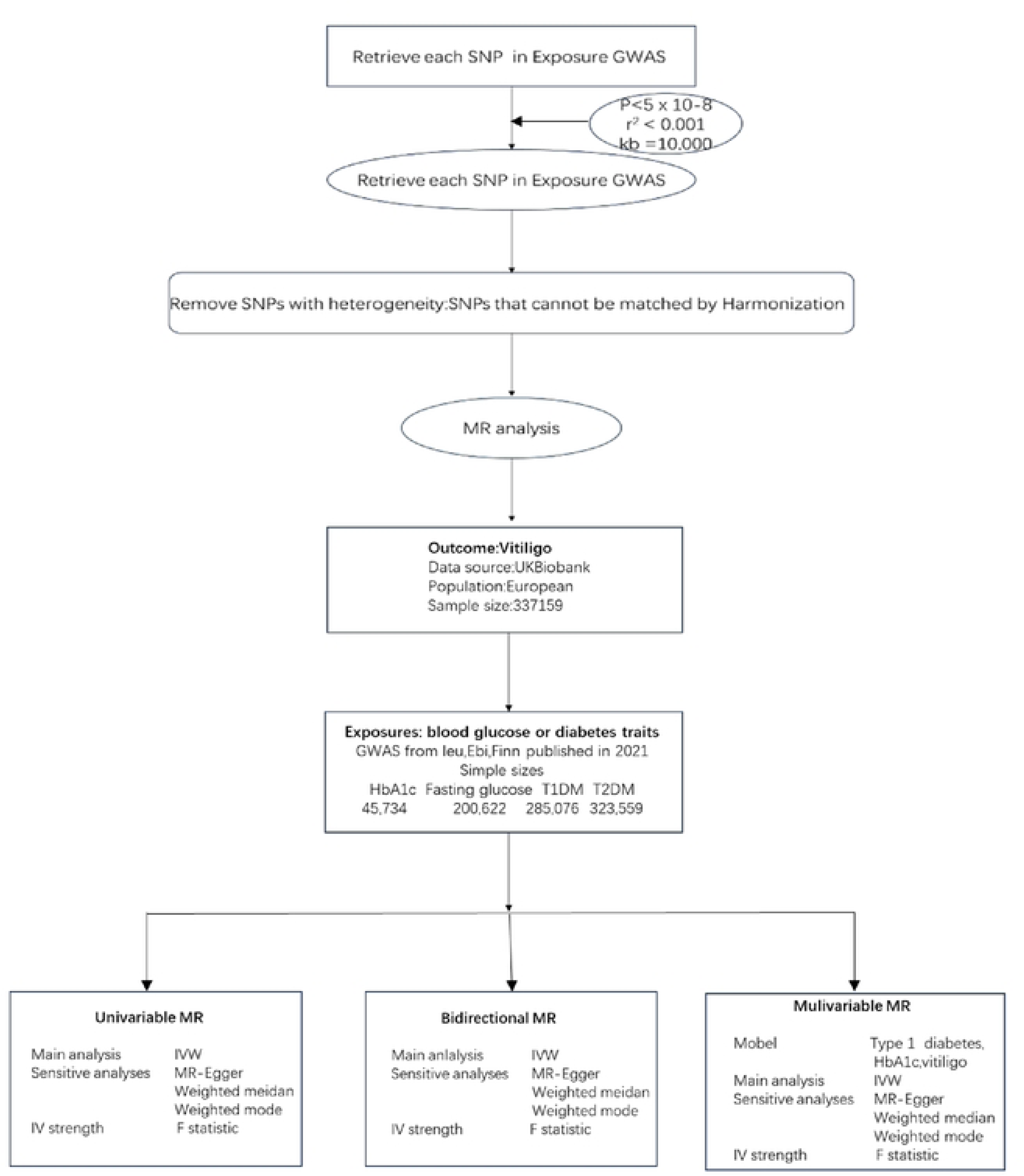
Study design and workflow.

**S2 Fig.**
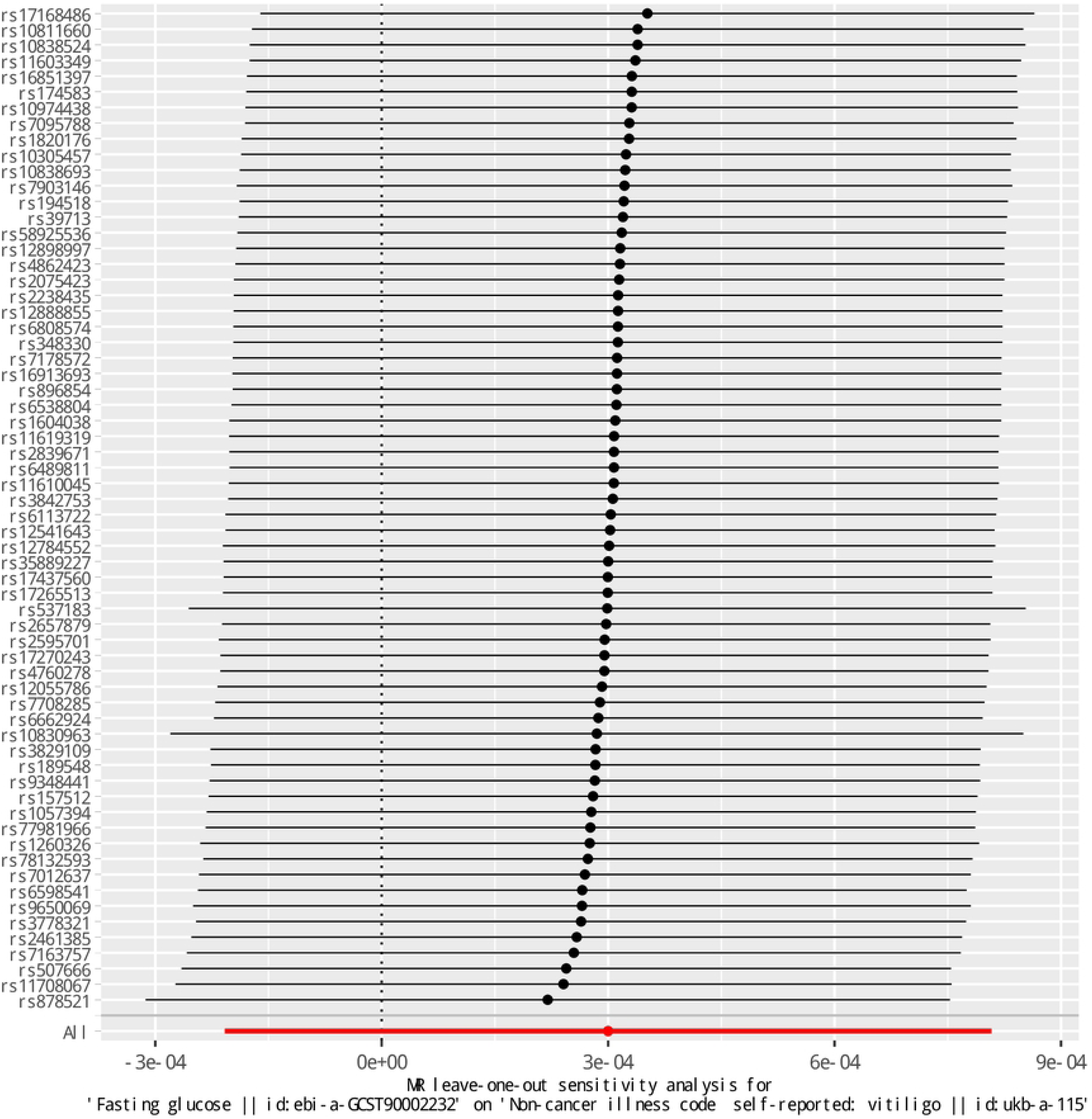
MR leave-one-out sensitivity analysis.

**S3 Fig.**
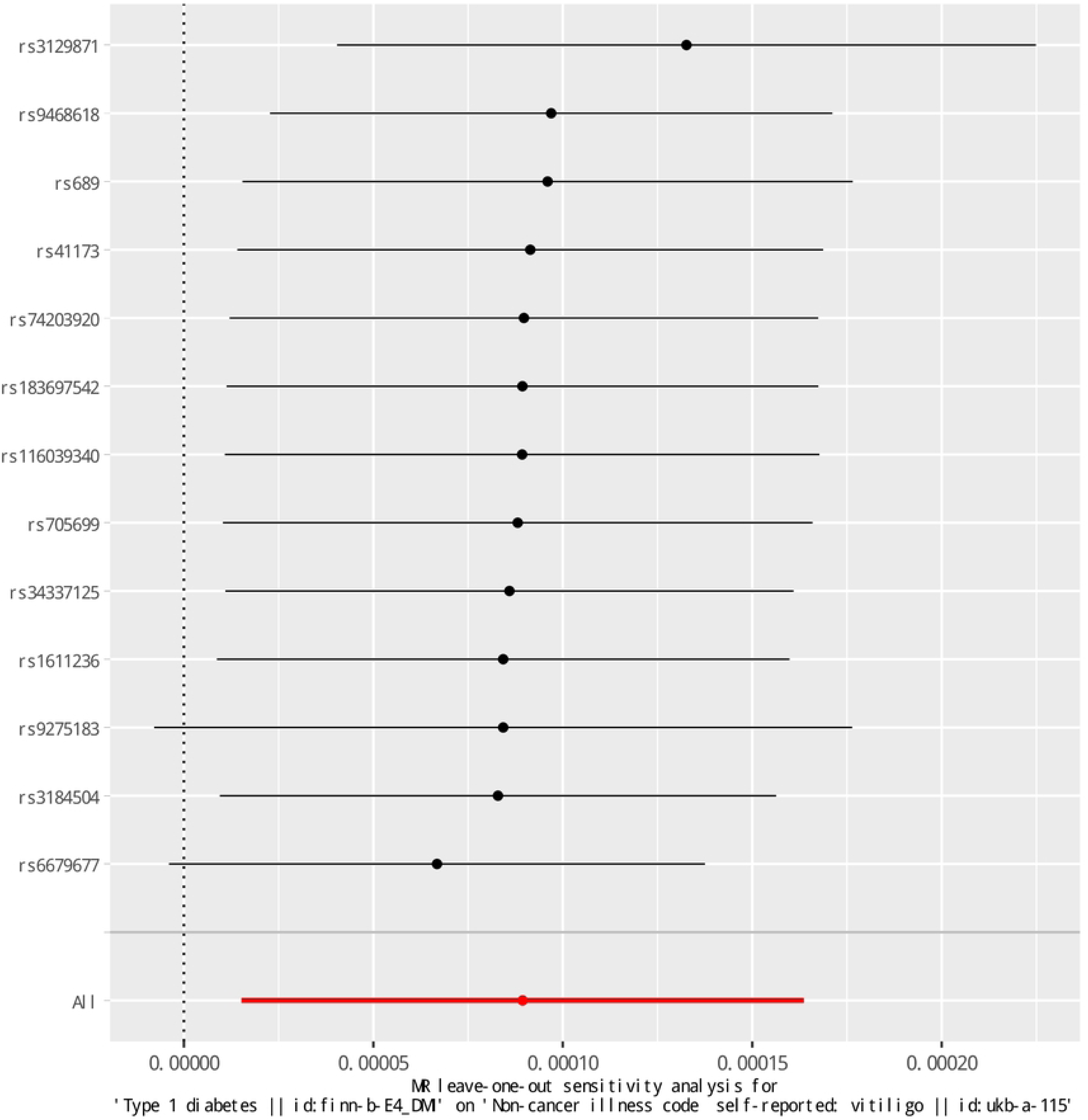
MR leave-one-out sensitivity analysis.

**S4 Fig.**
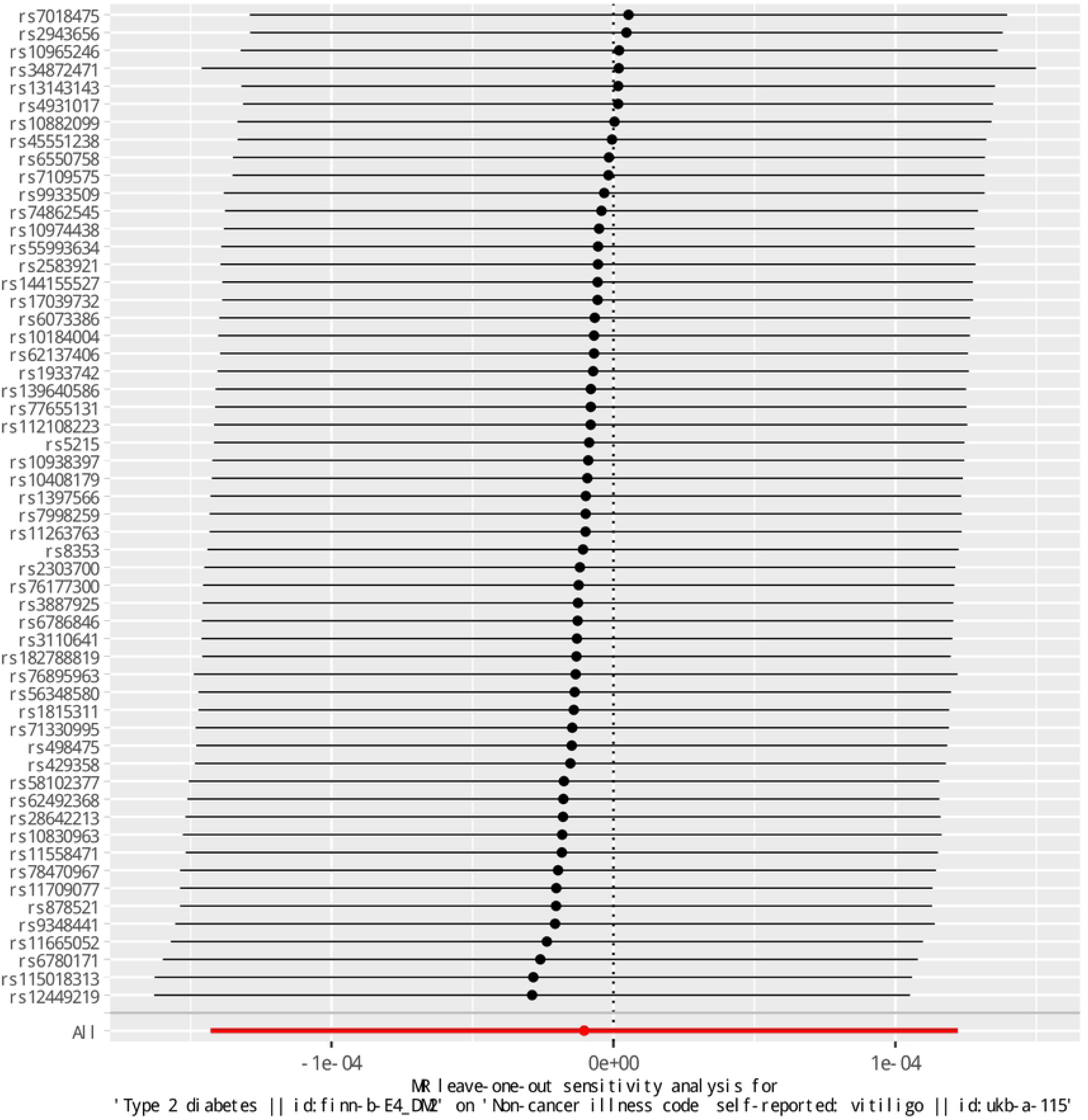
MR leave-one-out sensitivity analysis.

**S5 Fig.**
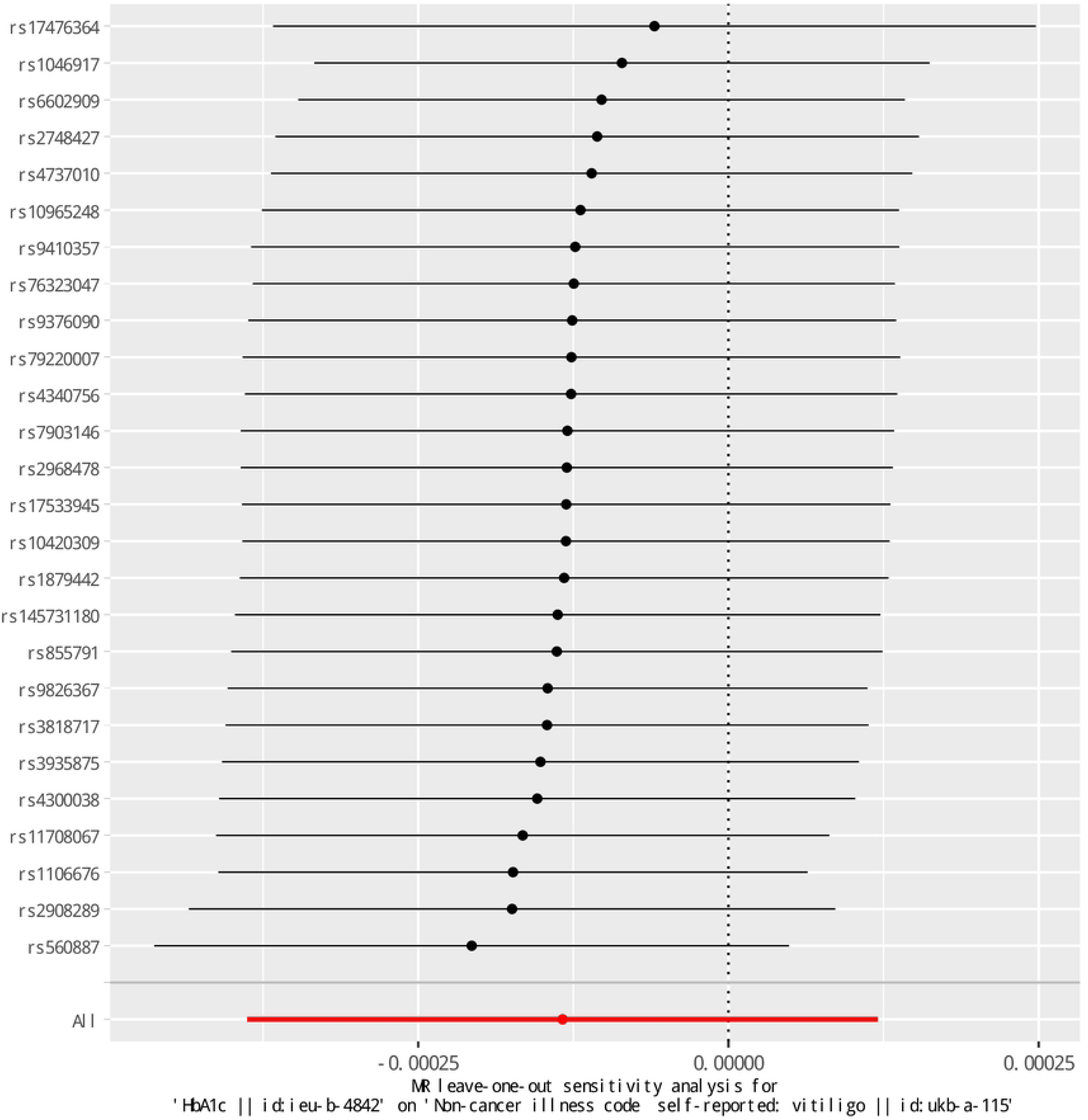
MR leave-one-out sensitivity analysis.

**S6 Fig.**
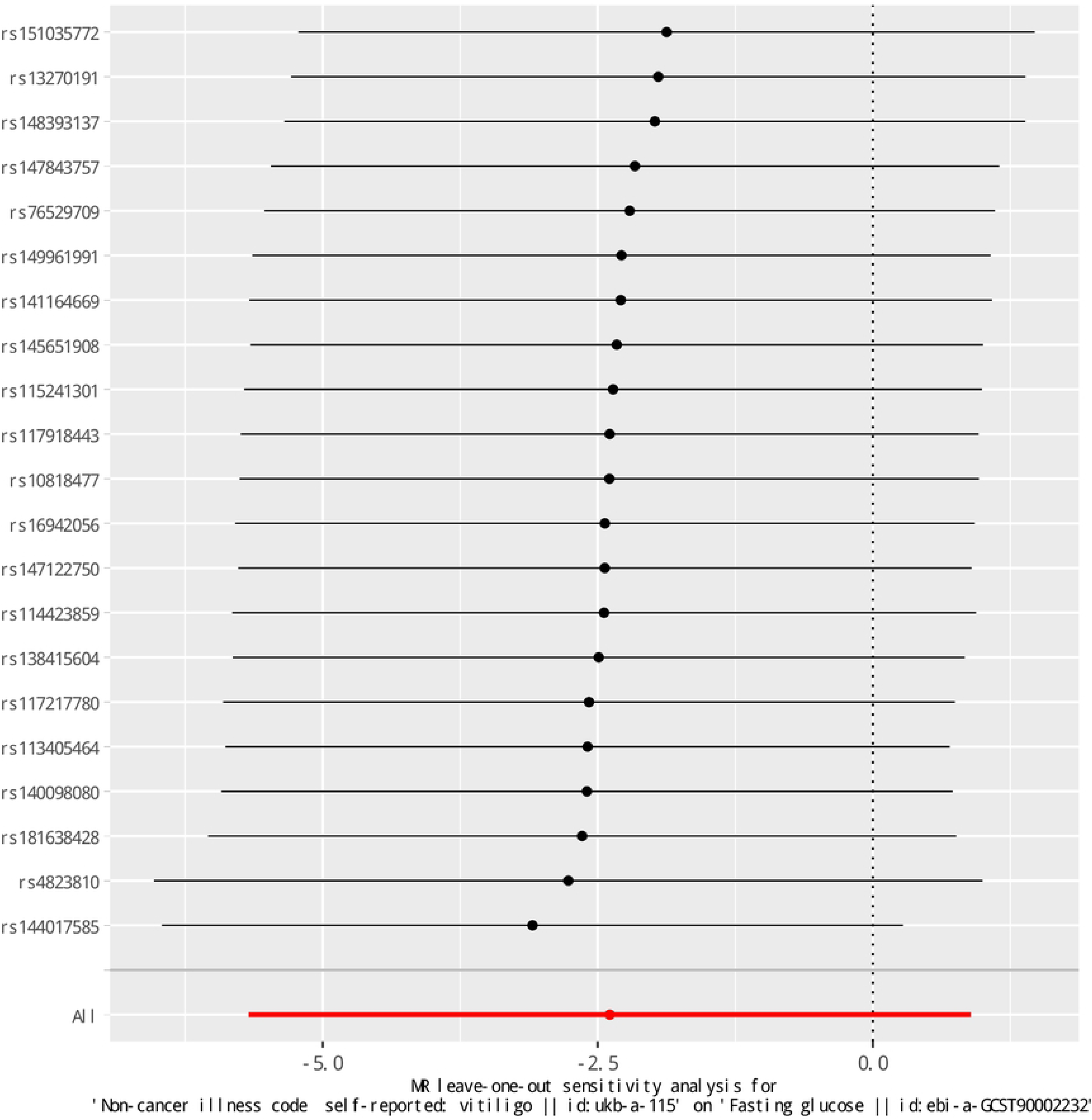
Reverse MR leave-one-out sensitivity analysis.

**S7 Fig.**
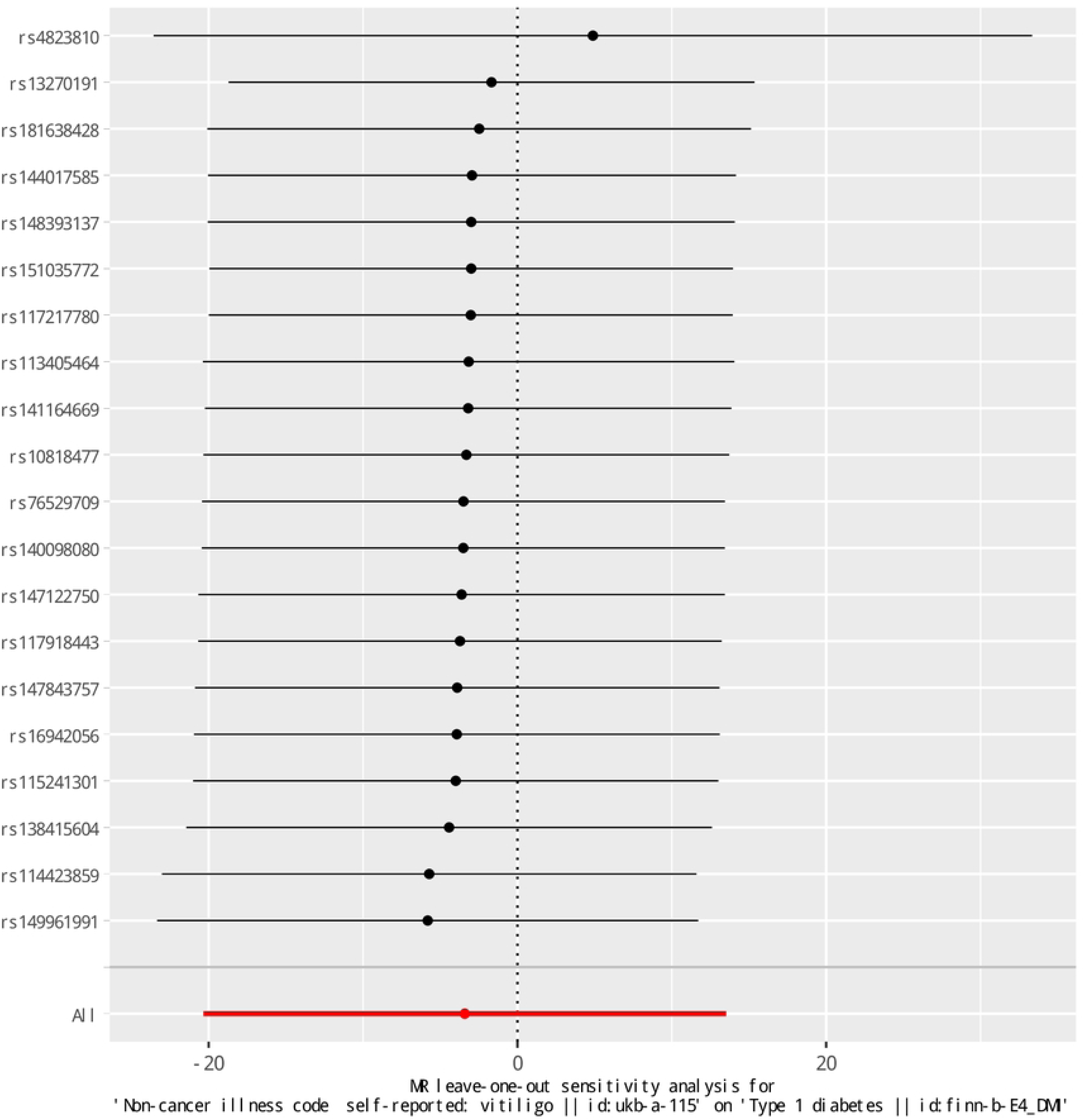
Reverse MR leave-one-out sensitivity analysis.

**S8 Fig.**
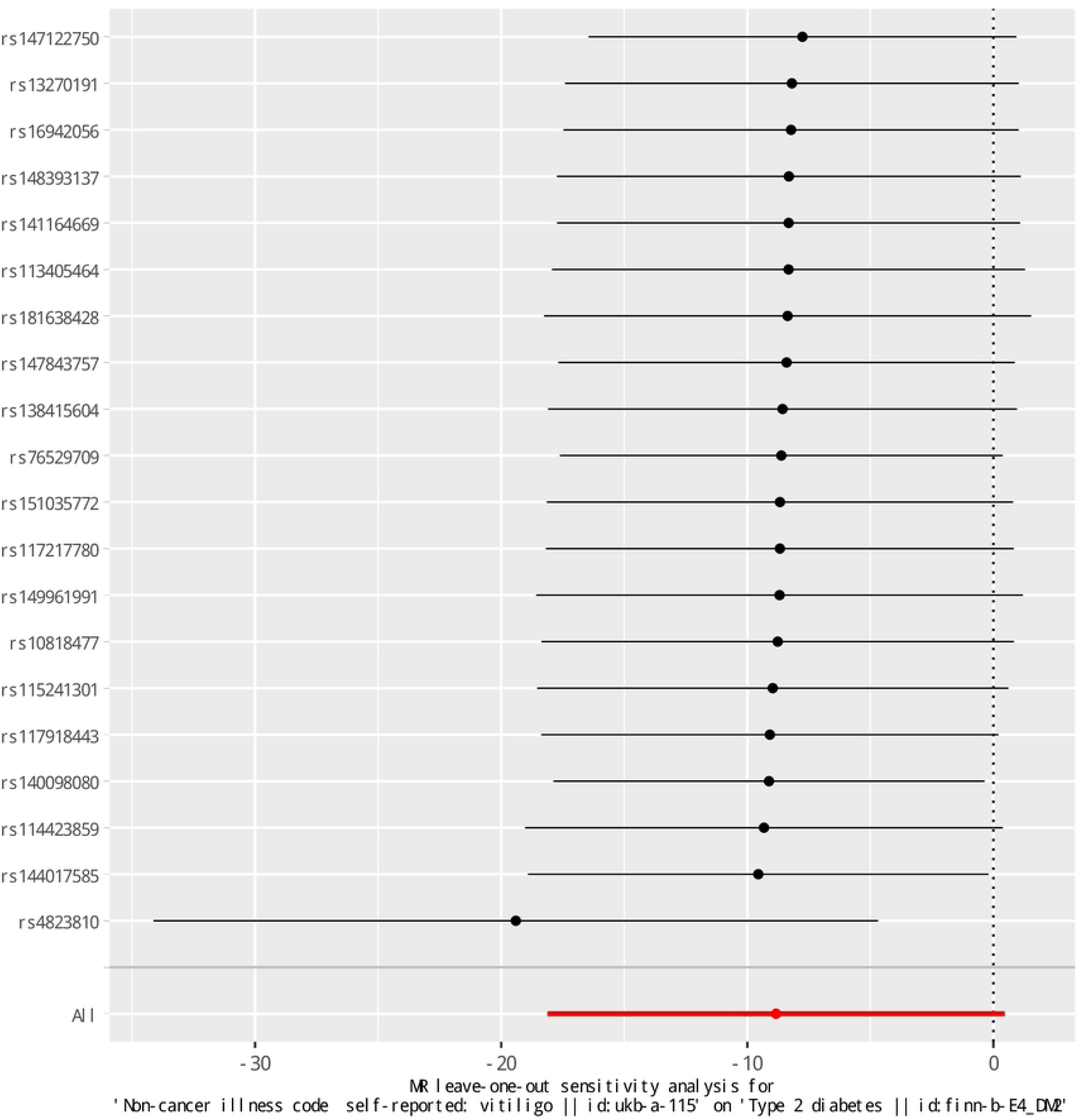
Reverse MR leave-one-out sensitivity analysis.

**S9 Fig.**
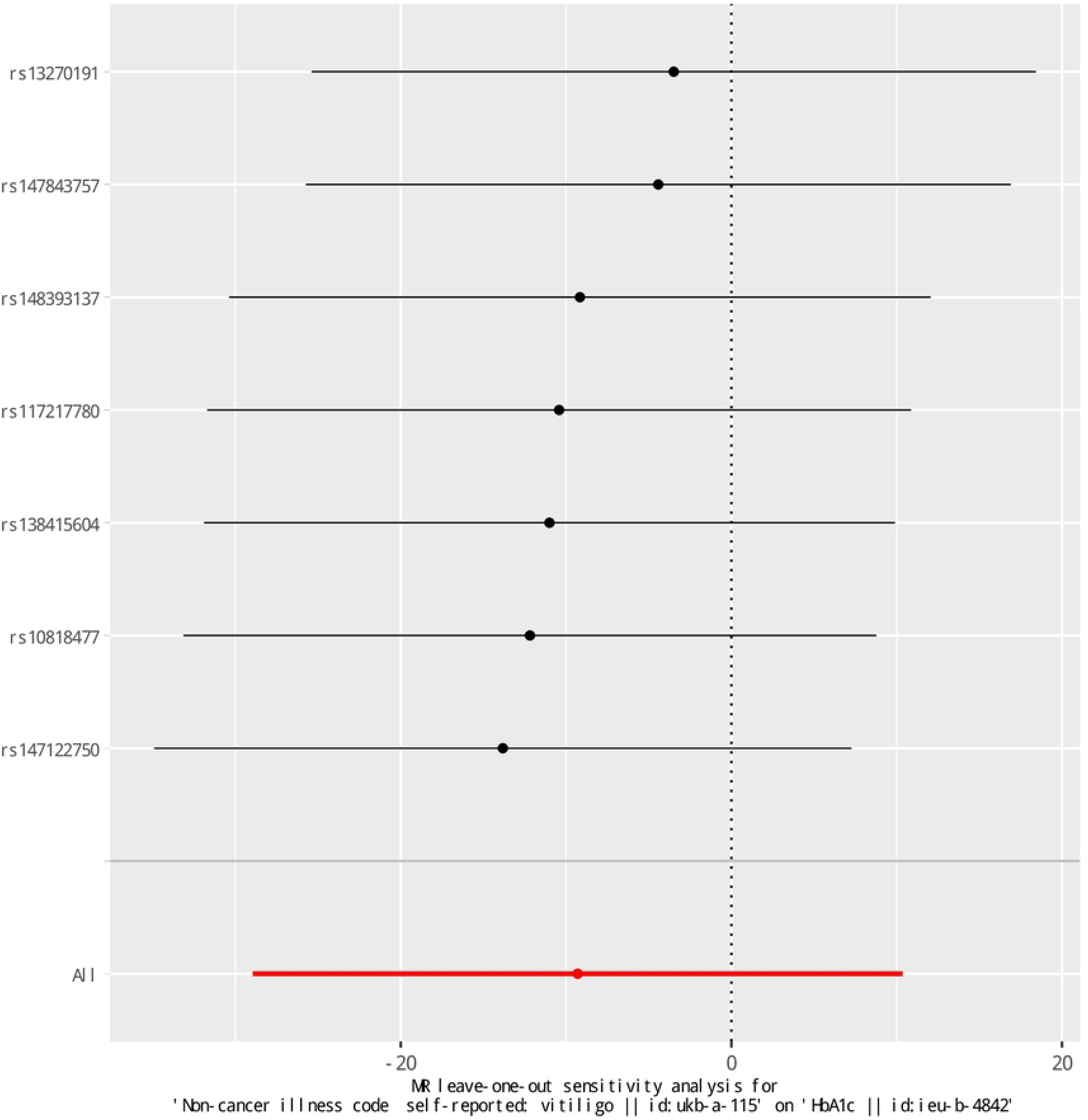
Reverse MR leave-one-out sensitivity analysis.

## Notes

### Competing Interest Statement

The authors have declared no competing interest.

### Author Declarations

We used aggregated genome-wide association data from the (Integrative Epidemiology Unit) IEU online database of European adults vitiligo; Glycated hemoglobin (HbA1c) data were from (IEU). Fasting blood glucose data were obtained from the European Bioinformatics Institute(EBI). T1DM and T2DM data were from FinnGen(FINN).

## References.

1. Alikhan A, Felsten LM, Daly M, Petronic-Rosic V. Vitiligo: a comprehensive overview Part I. Introduction, epidemiology, quality of life, diagnosis, differential diagnosis, associations, histopathology, etiology, and work-up. J Am Acad Dermatol. 2011;65(3):473–91.

2. Afkhami-Ardekani M, Ghadiri-Anari A, Ebrahimzadeh-Ardakani M, Zaji N. Prevalence of vitiligo among type 2 diabetic patients in an Iranian population. Int J Dermatol. 2014;53(8):956–8.

3. Chang HC, Lin MH, Huang YC, Hou TY. The association between vitiligo and diabetes mellitus: A systematic review and meta-analysis. J Am Acad Dermatol. 2019;81(6):1442–5.

4. Hosseini E, Mokhtari Z, Salehi Abargouei A, Mishra GD, Amani R. Maternal circulating leptin, tumor necrosis factor-alpha, and interleukine-6 in association with gestational diabetes mellitus: a systematic review and meta-analysis. Gynecol Endocrinol. 2023;39(1):2183049.

5. Burgess S, Thompson SG. Multivariable Mendelian randomization: the use of pleiotropic genetic variants to estimate causal effects. Am J Epidemiol. 2015;181(4):251–60.

6. Katan MB. Apolipoprotein E isoforms, serum cholesterol, and cancer. Lancet. 1986;1(8479):507-8.

7. Davey Smith G, Ebrahim S. What can mendelian randomisation tell us about modifiable behavioural and environmental exposures? Bmj. 2005;330(7499):1076- 9

8. Skrivankova VW, Richmond RC, Woolf BAR, Yarmolinsky J, Davies NM, Swanson SA, et al. Strengthening the Reporting of Observational Studies in Epidemiology Using Mendelian Randomization: The STROBE-MR Statement. Jama. 2021;326(16):1614–21.

9. Gough A, Sitch A, Ferris E, Marshall T. Within-subject variation of HbA1c: A systematic review and meta-analysis. PLoS One. 2023;18(8):e0289085.

10. Ghosh K, Das K, Ghosh S, Chakraborty S, Jatua SK, Bhattacharya A, et al. Prevalence of Skin Changes in Diabetes Mellitus and its Correlation with Internal Diseases: A Single Center Observational Study. Indian J Dermatol. 2015;60(5):465–9.

11. Kordonouri O, Maguire AM, Knip M, Schober E, Lorini R, Holl RW, et al. Other complications and associated conditions with diabetes in children and adolescents. Pediatr Diabetes. 2009;10 Suppl 12:204–10.

12. Greenland S. An introduction to instrumental variables for epidemiologists. Int J Epidemiol. 2000;29(4):722–9.

13. Burgess S, Thompson SG. Avoiding bias from weak instruments in Mendelian randomization studies. Int J Epidemiol. 2011;40(3):755–64.

14. Burgess S, Thompson SG. Interpreting findings from Mendelian randomization using the MR-Egger method. Eur J Epidemiol. 2017;32(5):377–89.

15. Bowden J, Davey Smith G, Haycock PC, Burgess S. Consistent Estimation in Mendelian Randomization with Some Invalid Instruments Using a Weighted Median Estimator. Genet Epidemiol. 2016;40(4):304–14.

16. Bowden J, Del Greco MF, Minelli C, Davey Smith G, Sheehan N, Thompson J. A framework for the investigation of pleiotropy in two-sample summary data Mendelian randomization. Stat Med. 2017;36(11):1783–802.

17. Nikolakopoulou A, Mavridis D, Salanti G. How to interpret meta-analysis models: fixed effect and random effects meta-analyses. Evid Based Ment Health. 2014;17(2):64.

18. Verbanck M, Chen CY, Neale B, Do R. Detection of widespread horizontal pleiotropy in causal relationships inferred from Mendelian randomization between complex traits and diseases. Nat Genet. 2018;50(5):693–8.

19. Paaby AB, Rockman MV. The many faces of pleiotropy. Trends Genet. 2013;29(2):66–73.

20. Kang P, Zhang WG, Ji ZH, Shao ZJ, Li CY. Association between vitiligo and relevant components of metabolic syndrome: a systematic review and meta-analysis. J Dtsch Dermatol Ges. 2022;20(5):629–41.

21. Yuan S, Merino J, Larsson SC. Causal factors underlying diabetes risk informed by Mendelian randomisation analysis: evidence, opportunities and challenges. Diabetologia. 2023;66(5):800–12.

22. Possidente C, Fanelli G, Serretti A, Fabbri C. Clinical insights into the cross-link between mood disorders and type 2 diabetes: A review of longitudinal studies and Mendelian randomisation analyses. Neurosci Biobehav Rev. 2023;152:105298.

23. Lai YC, Yew YW, Kennedy C, Schwartz RA. Vitiligo and depression: a systematic review and meta-analysis of observational studies. Br J Dermatol. 2017;177(3):708–18.

24. Yue Y, Tang Y, Tang J, Shi J, Zhu T, Huang J, et al. Maternal infection during pregnancy and type 1 diabetes mellitus in offspring: a systematic review and meta- analysis. Epidemiol Infect. 2018;146(16):2131–8.

25. Felton JL, Conway H, Bonami RH. B Quiet: Autoantigen-Specific Strategies to Silence Raucous B Lymphocytes and Halt Cross-Talk with T Cells in Type 1 Diabetes. Biomedicines. 2021;9(1).

26. Krischer JP, Lynch KF, Schatz DA, Ilonen J, Lernmark Å, Hagopian WA, et al. The 6 year incidence of diabetes-associated autoantibodies in genetically at-risk children: the TEDDY study. Diabetologia. 2015;58(5):980–7.

27. Oling V, Reijonen H, Simell O, Knip M, Ilonen J. Autoantigen-specific memory CD4+ T cells are prevalent early in progression to Type 1 diabetes. Cell Immunol. 2012;273(2):133–9.

28. Roep BO, Peakman M. Antigen targets of type 1 diabetes autoimmunity. Cold Spring Harb Perspect Med. 2012;2(4):a007781.

29. Frisoli ML, Essien K, Harris JE. Vitiligo: Mechanisms of Pathogenesis and Treatment. Annu Rev Immunol. 2020;38:621–48.

30. Rashighi M, Agarwal P, Richmond JM, Harris TH, Dresser K, Su MW, et al. CXCL10 is critical for the progression and maintenance of depigmentation in a mouse model of vitiligo. Sci Transl Med. 2014;6(223):223ra23.

31. Richmond JM, Masterjohn E, Chu R, Tedstone J, Youd ME, Harris JE. CXCR3 Depleting Antibodies Prevent and Reverse Vitiligo in Mice. J Invest Dermatol. 2017;137(4):982–5.

32. Edwards J, Wilmott JS, Madore J, Gide TN, Quek C, Tasker A, et al. CD103(+) Tumor-Resident CD8(+) T Cells Are Associated with Improved Survival in Immunotherapy-Naïve Melanoma Patients and Expand Significantly During Anti-PD-1 Treatment. Clin Cancer Res. 2018;24(13):3036–45.

33. Gould IM, Gray RS, Urbaniak SJ, Elton RA, Duncan LJ. Vitiligo in diabetes mellitus. Br J Dermatol. 1985;113(2):153–5.

34. Ibrahim S, El-Tahlawi S, Mogawer RM, El Ansary M, Esmat S, El-Hawary M. Different vitiligo characteristics as predictors of increased risk of metabolic syndrome and insulin resistance: A case-control study. J Cosmet Dermatol. 2022;21(12):7170–7.

